# Psychosis spectrum symptoms among individuals with schizophrenia-associated copy number variants and evidence of cerebellar correlates of symptom severity

**DOI:** 10.1101/2022.03.03.22271848

**Authors:** Esra Sefik, Ryan M. Guest, Katrina Aberizk, Roberto Espana, Katrina Goines, Derek M. Novacek, Melissa M. Murphy, Adam E. Goldman-Yassen, Joseph F. Cubells, Opal Ousley, Longchuan Li, Sarah Shultz, Elaine F. Walker, Jennifer G. Mulle

## Abstract

The 3q29 deletion (3q29Del) is a copy number variant (CNV) with one of the highest effect sizes for psychosis-risk (>40-fold). Systematic research offers promising avenues for elucidating mechanism; however, compared to CNVs like 22q11.2Del, 3q29Del remains understudied. Emerging findings indicate that posterior fossa abnormalities are common among carriers, but their clinical relevance is unclear. Here, we report the first in-depth evaluation of psychotic symptoms in participants with 3q29Del (*N*=23), using the Structured Interview for Psychosis-Risk Syndromes, and compare this profile to 22q11.2Del (*N*=31) and healthy controls (*N*=279). By neuroimaging, we also explore correlations between psychotic symptoms and posterior fossa abnormalities in 3q29Del. Cumulatively, 48% of the 3q29Del sample exhibited a psychotic disorder or attenuated positive symptoms, with a subset meeting criteria for clinical high-risk. 3q29Del had more severe ratings than controls on all domains and only exhibited less severe ratings than 22q11.2Del in negative symptoms, with select sex differences. An inverse relationship was identified between positive symptoms and cerebellar cortex volume in 3q29Del, documenting the first clinically-relevant neuroanatomical connection in this syndrome. Our findings characterize the profile of psychotic symptoms in the largest 3q29Del sample reported to date, contrast with another high-impact CNV, and highlight cerebellar involvement in elevated psychosis-risk.

**Highlights:**

- The rare copy number variants (CNVs) 3q29Del and 22q11.2Del confer the largest known effect sizes for schizophrenia susceptibility.
- While 22q11.2Del’s link with psychosis has been extensively studied, in-depth characterization of psychotic symptoms associated with 3q29Del is lacking.
- This study fills this gap and provides the first phenotypic comparison of psychotic symptom profiles in carriers of these CNVs, and healthy controls.
- Additionally, we uncover a novel genetic association between 3q29Del, psychotic symptoms, and cerebellar cortex development.

## 1. Introduction

The identification of copy number variants (CNVs) associated with high risk for serious mental illnesses has opened new avenues for investigating etiology. Two such CNVs, the 3q29 and 22q11.2 deletions (Del), significantly increase risk for a range of neuropsychiatric disorders, especially schizophrenia and other psychoses (Malhotra and Sebat, 2012; Marshall et al., 2017; Mulle, 2015; Rees et al., 2014; Schneider et al., 2014; Stefansson et al., 2008). Systematic research on these CNVs holds promise for shedding light on the neuropathology underlying psychosis.

22q11.2Del is caused by a hemizygous 1.5- to 3.0-Mb deletion involving 45 protein-coding genes. The deletion affects ∼1 in 2,148-4,000 individuals (Blagojevic et al., 2021; Botto et al., 2003; Mcdonald-Mcginn et al., 2001) and conveys a >20-fold increased risk for schizophrenia (Fung et al., 2010; McDonald-McGinn et al., 2015). Standardized measures have been used to examine the clinical high-risk (CHR) or “prodromal” symptoms that may precede the onset of psychosis in 22q11.2Del syndrome and the general population. Before their first full-blown psychotic episode, typically during late-adolescence and early-adulthood, individuals often exhibit a period of functional decline coinciding with the emergence of attenuated psychotic symptoms (Goulding et al., 2013; Vorstman et al., 2015; Yung and McGorry, 1996). Those identified at CHR present with these “warning” signs and are at substantial risk for future psychosis (Salazar de Pablo et al., 2021). Reports in 22q11.2Del have shown that about 30-60% of study participants manifest attenuated psychotic symptoms, which are known to be linked with elevated risk for subsequent conversion to psychosis (Shapiro et al., 2011; Weisman et al., 2017).

Individuals with 3q29Del are hemizygous for a 1.6-Mb interval containing 21 protein-coding genes. Compared to 22q11.2Del, 3q29Del was only recently identified (Cox and Butler, 2015) and is less common, with a prevalence of ∼1 in 30,000 individuals (Sanchez Russo et al., 2021); consequently the associated phenotypes are just now being documented. Like 22q11.2Del, 3q29Del is pleiotropic, elevating risk for a range of physical and psychiatric disabilities, including cardiac defects, cognitive deficits, autism spectrum disorder, and attention-deficit/hyperactivity disorder (Glassford et al., 2016; Pollak et al., 2019). Current evidence suggests that 3q29Del confers an equal or greater risk for psychosis than 22q11.2Del. Recent reports indicate that 3q29Del confers a >40-fold increased risk for schizophrenia over the general population (Marshall et al., 2017; Mulle, 2015; Singh et al., 2020). It is not known whether early clinical signs of psychosis risk at various stages of development are elevated among individuals with 3q29Del.

Our team recently documented the overarching neurodevelopmental and psychiatric manifestations associated with 3q29Del through direct evaluation and magnetic resonance imaging (MRI) (Sanchez Russo et al., 2021). Here, we report results on psychotic symptoms in study participants with 3q29Del, utilizing the gold-standard Structured Interview for Psychosis-Risk Syndromes (SIPS) (Miller et al., 2003). We examine the proportion of 3q29Del participants with attenuated or florid psychotic symptoms, incorporate both categorical and dimensional elements of evaluation, and contrast these findings with the symptom profile in 22q11.2Del and healthy controls (HC).

Additionally, we explore the association between psychosis susceptibility in 3q29Del and structural brain imaging findings. Our recent MRI research discovered a striking increase in cerebellar hypoplasia and cystic/cyst-like malformations of the posterior fossa in individuals with 3q29Del (Sanchez Russo et al., 2021), with a frequency that surpasses all other radiological anomalies documented in this population (Sefik et al., 2022). Moreover, we have shown quantitatively that cerebellar structure differs volumetrically from HCs (Sefik et al., 2022). These findings coincide with emerging literature on cerebellar contributions to psychiatric disorders, and the therapeutic potential of cerebellar neuromodulation in various diseases, however their clinical implications in 3q29Del are unknown. Given the pronounced nature of posterior fossa abnormalities within this population, we center our focus on this region and explore its relevance to elevated psychosis risk in one of the highest-impact CNVs associated with schizophrenia.

## 2. Methods

### 2.1. Participants

#### 3q29Del sample

Twenty-three individuals with 3q29Del, ages 8.08-39.12 (mean ± SD = 16.94 ± 8.24 years) were evaluated using the SIPS. Participants were ascertained from the 3q29 registry (Glassford et al., 2016) and between August 2017-February 2020 traveled to Atlanta, GA for deep phenotyping and MRI. Deletion status was confirmed from clinical genetics and/or medical records. The project protocol, sample characteristics and comorbidities have been described (Murphy et al., 2018; Sanchez Russo et al., 2021).

#### Comparison samples

The HC sample included 279 individuals, ages 12.07-34.41 (mean ± SD = 20.08 ± 4.61 years), from the second cohort of the North American Prodromal Longitudinal Study (NAPLS2), who were evaluated with the SIPS between June 2009-October 2012 (Addington et al., 2015). The HCs neither met criteria for any psychotic disorder nor reported attenuated psychotic symptoms, and had no central nervous system disorder, intellectual disability, substance dependence in the previous six months, or a first-degree relative with a psychotic disorder.

The 22q11.2Del sample included SIPS data from 31 individuals, ages 13.85-29.77 (mean ± SD = 19.75 ± 4.12 years), from the 22q11.2 Clinic in Atlanta, GA (Rockers et al., 2009; Shapiro et al., 2011; Weisman et al., 2017). Participants were ascertained from a case registry, with the deletion confirmed via fluorescence in situ hybridization (FISH). Participants were often referred for FISH as minors due to cardiovascular defects, language difficulties or immunological problems. Individuals identified as adults were referred as part of clinical care within a genetics or heart clinic (Shapiro et al., 2011). Data were collected between February 2006-June 2007.

### 2.2. Assessment of attenuated and florid psychotic symptoms

All participants were administered the SIPS by trained personnel. Raters of both CNV samples were supervised by the same psychologist (E.F.W.). The 19 items in the semi-structured interview are grouped within four domains/constructs: positive, negative, disorganization, general. Each item is rated from 0 (*Absent*) to 6 (*Severe and Psychotic* for positive symptoms, *Extreme* for others). Positive symptom ratings of 3-5 indicate “attenuated” psychotic symptoms, whereas 6 indicates “florid” psychotic symptoms that lead to disorganized or dangerous behaviors and a complete loss of insight. Item ratings were summed to produce a total score for each domain. For individuals meeting criteria for psychosis, the Structured Clinical Interview for DSM-5 (SCID-5) (First et al., 2015) or -4 (SCID-4) (First and Gibbon, 2004) specified the diagnosis. Studies were approved by appropriate institutional review boards. All participants provided informed consent.

### 2.3. MRI data acquisition and processing

T1- and T2-weighted structural MRI was performed in 17 medically eligible 3q29Del participants with SIPS data, ages 8.08-39.12 (mean ± SD = 18.13 ± 9.03 years). Images were acquired on a Siemens Magnetom Prisma 3T scanner in the sagittal plane using a 32-channel Prisma head coil and an 80mT/m gradient. T1- weighted 3D images were acquired using a single-echo MPRAGE sequence: TE=2.24ms, TR=2400ms, TI=1000ms, bandwidth=210Hz/pixel, FOV=256x256mm, resolution=0.8mm isotropic. T2-weighted 3D images were acquired using a SPACE sequence: TE=563ms, TR=3200ms, bandwidth=745Hz/pixel, FOV=256x256mm, resolution=0.8mm isotropic. Further details have been described (Sanchez Russo et al., 2021; Sefik et al., 2022). All images were processed with the “minimal structural pre-processing” pipeline established by the Human Connectome Project (v.4.1.3) using FreeSurfer (v.6.0) (http://surfer.nmr.mgh.harvard.edu/). Cerebellar segmentations were performed automatically as described in (Glasser et al., 2013), leveraging a probabilistic atlas and Bayesian classification rule that assigns anatomical labels to each voxel based on estimates derived from a manually labeled training set (Fischl et al., 2002). This yielded tissue-type specific volumes for cerebellar cortex and white matter, which were additionally summed to derive a global metric for total cerebellar volume, allowing methodological consistency with previous literature. Estimated total intracranial volume (eICV) was calculated by FreeSurfer (Buckner et al., 2004), and used as a proxy for head size. We report results from both eICV-adjusted and unadjusted analyses, given reports of microcephaly (Ballif et al., 2008; Cox and Butler, 2015; Willatt et al., 2005), and eICV reduction in individuals with 3q29Del relative to HCs (Sefik et al., 2022). See Supplemental Materials for further details on the neuroimaging protocol and quality control of anatomical segmentations.

### 2.4. Statistical analyses

All analyses were conducted using *R* (v.4.0.3). Frequencies of florid or attenuated psychotic symptoms were compared between groups using Fisher’s exact test. To assess the variation in symptom profiles between groups, a series of analyses of covariance (ANCOVA) were performed; first, by domain and then item-wise. Pairwise contrasts between 3q29Del and the two other groups were performed using Tukey’s test and corrected for multiple comparisons, using a *p-value* threshold of 0.01 for each SIPS domain (0.05/4 domains) and 0.003 for individual SIPS items (0.05/19 items). To address violations of ANCOVA assumptions (partially due to a floor effect observed in HCs), SIPS scores were log-transformed in between-group analyses.

For neuroimaging analyses, we used multiple linear regression to examine the relationship between cerebellar volumes and SIPS ratings for positive, negative and disorganization symptom domains among 3q29Del participants. We additionally corrected for eICV to test whether results reflect a link with the cerebellum beyond global variability in head size. We also employed a subsequent categorical approach, using binary logistic regression to estimate the probability that either a psychotic disorder or attenuated psychotic symptom syndrome (APSS) diagnosis is present in the 3q29Del sample given cerebellar volumes as the predictor. Multiple comparisons correction was applied at the ROI level, using a *p-value* threshold of 0.02 (0.05/3 ROIs).

Finally, we tested whether psychotic symptoms differ between 3q29Del participants with versus without cystic/cyst-like malformations of the posterior fossa, identified by a board-certified neuroradiologist (Sefik et al., 2022). Fisher’s exact test was used for diagnostic comparisons. Student’s t-test and Wilcoxon signed-rank test were used for dimensional comparisons. Given limited power due to smaller sample size in this arm, we also report trend-level associations using a less conservative alpha (*p-value* ≤0.10) to inform future hypotheses.

All analyses were two-tailed. Age and sex were considered as covariates. Given observed violations of linear regression assumptions using ordinary least squares, heteroscedasticity-robust estimates (estimation-type: HC1) (MacKinnon and White, 1985) were calculated. See Supplemental Materials for extended methods.

## 3. Results

### 3.1. Presence of attenuated and florid psychotic symptoms in 3q29Del and 22q11.2Del

Demographics are presented in Table 1. Four out of 23 3q29Del participants (17%) met criteria for a psychotic disorder: schizophrenia (*N*=1); schizoaffective disorder, bipolar type (*N*=1); and unspecified schizophrenia spectrum and other psychotic disorder (*N*=2). Two had a prior diagnosis of a psychotic disorder; two did not. These individuals tended to be older than those without a psychotic disorder. However, cognitive ability was similar between the groups. Participants meeting criteria for a psychotic disorder reported past or current symptoms that met a rating of *Severe and Psychotic* for unusual thought content, suspiciousness, and/or perceptual abnormalities, and reported prodromal symptoms beginning in late-childhood or early-adolescence (ages 11-12 on average). In contrast, only one out of 31 22q11.2Del participants (3%) met criteria for a psychotic disorder.

**Table 1.** Demographic and clinical information for each sample and comparison with 3q29Del. . Descriptive statistics are reported separately for the total 3q29Del sample, and the sample stratified by presence/absence of a psychotic disorder. For race and ethnicity, 22q11.2Del *N*=17 and *N*=19, respectively. For general cognitive abilities, HC *N*=262. Percentages reflect fraction of participants with complete data. *^a^*Wilcoxon rank sum test, *^b^*Pearson’s chi-squared test. *^c^*Fisher’s exact test (due to smaller subsamples). *p-value* ≤0.001‘***’, *p-value* ≤0.01‘**’, *p-value ≤0.05‘*’*, *p-value* ≤0.10‘^†^’. *Abbreviations: healthy control, HC; standard deviation, SD*.

|  | 3q29Del<br>(N=23) |  |  | HC<br>(N=279) | 22q11.2Del<br>(N=31) | Pairwise comparisons |  |
| --- | --- | --- | --- | --- | --- | --- | --- |
|  | 3q29Del<br>(N=23) | With psychotic<br>disorder (N=4) | Without psychotic<br>disorder (N=19) |  |  | 3q29Del vs.<br>HC | 3q29Del vs.<br>22q11.2Del |
| **Age (years)** |  |  |  |  |  |  |  |
| Mean ± SD | 16.94 ± 8.24 | 26.66 ± 9.55 | 14.89 ± 6.51 | 20.08 ± 4.76 | 19.75 ± 4.12 | *p*-value<sup>a</sup> | *p*-value<sup>a</sup> |
| Median | 15.89 | 25.70 | 14.01 | 20.04 | 19.10 | = 0.003** | = 0.02* |
| Range | 8.08–39.12 | 16.13–39.12 | 8.08–34.64 | 12.07–34.41 | 13.85–29.77 |  |  |
| **Sex, N (%)** |  |  |  |  |  |  |  |
| Male | 14 (61%) | 3 (75%) | 11 (58%) | 141 (51%) | 14 (45%) | *p*-value<sup>b</sup> | *p*-value<sup>b</sup> |
| Female | 9 (39%) | 1 (25%) | 8 (42%) | 138 (49%) | 17 (55%) | = 0.46 | = 0.39 |
| **Race, N (%)** |  |  |  |  |  |  |  |
| White | 20 (87%) | 4 (100%) | 16 (84%) | 152 (55%) | 13 (76%) | *p*-value<sup>c</sup><br>= 0.006** | *p*-value<sup>c</sup><br>= 0.02* |
| Black or African American | 0 (0%) | 0 (0%) | 0 (0%) | 48 (17%) | 3 (18%) |  |  |
| Asian | 0 (0%) | 0 (0%) | 0 (0%) | 30 (11%) | 1 (6%) |  |  |
| More than one race | 3 (13%) | 0 (0%) | 3 (16%) | 29 (10%) | 0 (0%) |  |  |
| Other (i.e., Pacific Islander) | 0 (0%) | 0 (0%) | 0 (0%) | 20 (7%) | 0 (0%) |  |  |
| **Ethnicity, N (%)** |  |  |  |  |  |  |  |
| Hispanic/Latino | 1 (4%) | 0 (0%) | 1 (5%) | 50 (18%) | 2 (11%) | *p*-value<sup>c</sup> | *p*-value<sup>c</sup> |
| Non-Hispanic/Latino | 22 (96%) | 4 (100%) | 18 (95%) | 229 (82%) | 17 (89%) | = 0.14 | = 0.58 |
| **General cognitive abilities** |  |  |  |  |  |  |  |
| Mean ± SD | 73.74 ± 11.98 | 73.00 ± 14.72 | 73.89 ± 11.80 | 111.00 ± 14.16 | 73.71 ± 15.91 | *p*-value<sup>a</sup> | *p*-value<sup>a</sup> |
| Median | 75 | 76 | 76 | 112 | 71 | <0.001*** | = 0.88 |
| Range | 46–99 | 61–94 | 46–99 | 72–139 | 46–109 |  |  |

We next analyzed subthreshold psychotic symptoms among individuals without a psychotic disorder and with complete data (*N*=19 in 3q29Del, *N*=29 in 22q11.2Del). Seven 3q29Del participants (37%) exhibited one or more clinically significant attenuated positive symptoms (rated ≥3). Three of these individuals (16%) met the frequency and timing criteria for APSS (Miller et al., 2003). Nineteen 22q11.2Del participants (66%) exhibited clinically significant attenuated positive symptoms, which is at the higher end of rates found in 22q11.2Del samples (Weisman et al., 2017). The information necessary for assessing APSS was unavailable for the 22q11.2Del group.

In the full 3q29Del sample, the two most prevalent clinically significant attenuated positive symptoms were disorganized communication (30%) and perceptual abnormalities (30%), followed by unusual thought content (22%), suspiciousness (13%), and grandiosity (9%). In the 22q11.2Del sample, the two most prevalent clinically significant attenuated positive symptoms were suspiciousness (29%) and disorganized communication (23%), followed by perceptual abnormalities (19%), unusual thought content (13%) and grandiosity (13%). Cumulatively, 48% of the 3q29Del sample showed either florid or attenuated psychotic symptoms, as compared to 67% of the 22q11.2Del sample (Table 2, Fig. S1); the comparison of these frequencies revealed no significant difference between the groups (*p-values* >0.05).

**Table 2.** Rates of clinically significant psychotic symptoms (at least one item rated ≥3) among 3q29Del and 22q11.2Del participants. For P5 and the positive symptom domain total, 22q11.2Del *N*=30. *N*=13 3q29Del participants and *N*=10 22q11.2Del participants had >1 P-item rated ≥3.

|  | **3q29Del**<br>(N=23) | **22q11.2Del**<br>(N=31) |
| --- | --- | --- |
|  | Clinically significant,<br>N (%) | Clinically significant,<br>N (%) |
| **Positive Symptom Domain** | 11 (48%) | 20 (67%) |
| P1. Unusual Thought Content | 5 (22%) | 4 (13%) |
| P2. Suspiciousness | 3 (13%) | 9 (29%) |
| P3. Grandiosity | 2 (9%) | 4 (13%) |
| P4. Perceptual Abnormalities | 7 (30%) | 6 (19%) |
| P5. Disorganized Communication | 7 (30%) | 7 (23%) |

### 3.2. Group differences in demographics and symptom profiles

SIPS symptom ratings are presented in Table 3. There was a significant age difference between the 3q29Del sample and HCs (*p-value* ≤0.01), and between 3q29Del and 22q11.2Del (*p-value* ≤0.05), with 3q29Del participants being younger on average. Given these age differences, as well as evidence indicating increases in psychotic-like experiences through adolescence followed by decline in early-adulthood (Hafner, 2019; Sullivan et al., 2020; Waford et al., 2015), we first assessed whether the relationship between age and symptom severity differs between study samples. Results are presented in Table S1 and Fig. S2. Since the mean age and correlations between symptom severity and age differed between groups, our initial comparison of symptom profiles in the complete sample did not involve age adjustment. This approach was taken because covariate adjustment using ANCOVA assumes homogeneity of regression slopes across groups. To address possible confounding effects, we additionally performed supplemental analyses involving a smaller subset of participants matched for age and sex.

**Table 3.**
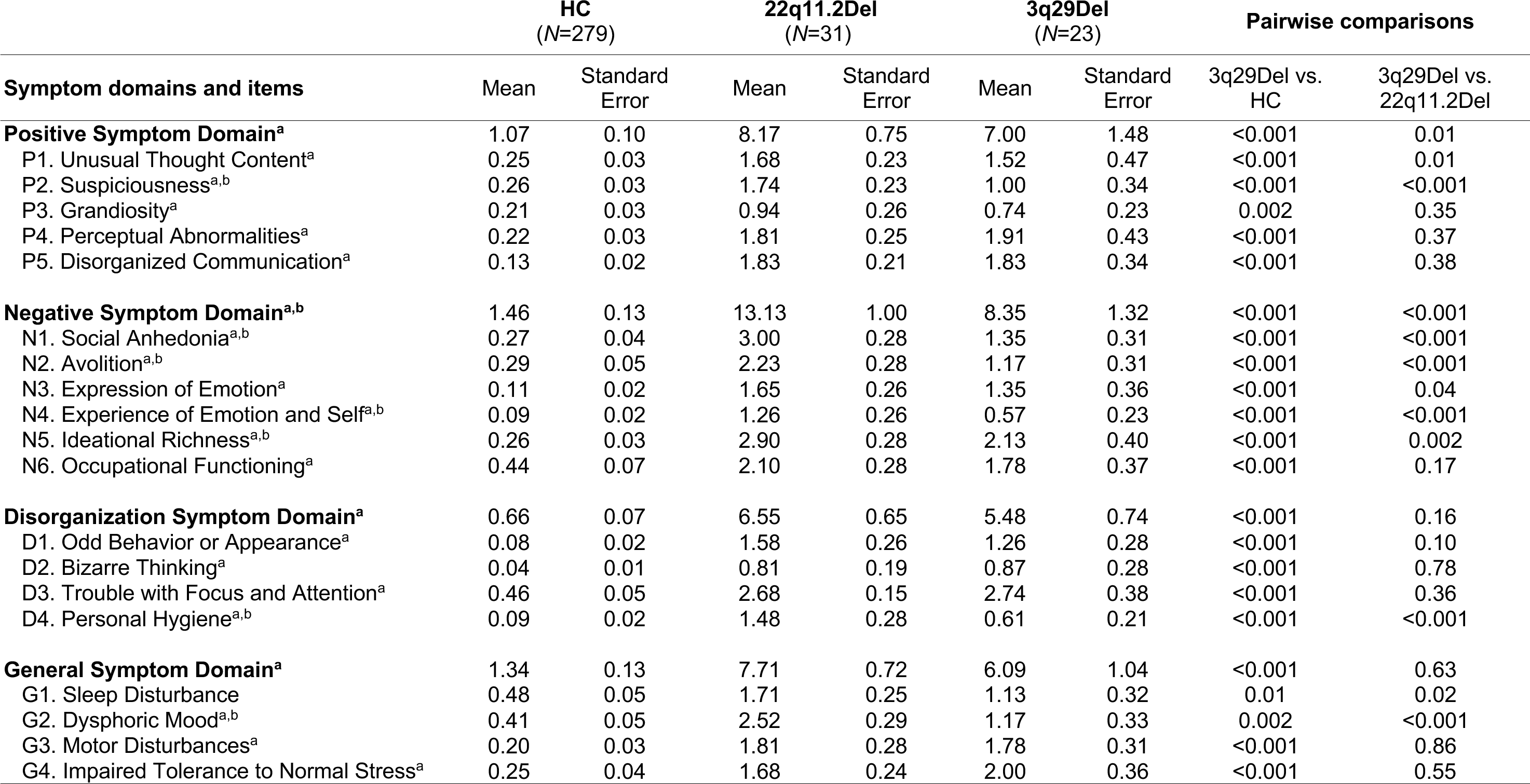
Overall results of the ANCOVA between SIPS ratings of each diagnostic group and pairwise comparisons. Unadjusted means and standard errors are shown. Pairwise comparisons reflect p-values calculated on log-transformed sex-adjusted data. ‘^a^’ indicates significant difference (*p-value* ≤0.01 for domain, *p- value* ≤0.003 for item) between 3q29Del and HC; ‘^b^’ indicates significant difference (*p-value* ≤0.01 for domain, *p- value* ≤0.003 for item) between 3q29Del and 22q11.2Del. For P5 and the positive symptom domain total, 22q11.2Del *N*=30. For G2 and the general symptom domain total, HC *N*=278. *Abbreviations:* Structured Interview for Psychosis-Risk Syndromes, SIPS; healthy controls, HC.

ANCOVAs performed in the complete sample contrasted the average symptom ratings by domain of the 3q29Del sample against HC and 22q11.2Del, adjusting for sex. There was a significant effect of diagnostic group on all domains: positive [*F*(2, 328) = 112.87, *p-value ≤*0.001, η^2^p = 0.41], negative [*F*(2, 329) = 135.00, *p-value*≤0.001, η^2^p = 0.45], disorganization [*F*(2, 329) = 166.30, *p-value* ≤0.001, η^2^p = 0.50], general [*F*(2, 328) = 74.56, *p-value* ≤0.001, η^2^p = 0.31]. Individuals with 3q29Del were rated significantly higher than HCs on all SIPS domains (*p-value*s ≤0.01): positive [*t*(328) = 8.12, *p-value* ≤0.001], negative [*t*(329) = 8.21, *p-value* ≤0.001], disorganization [*t*(329) = 11.10, *p-value* ≤0.001], and general [*t*(328) = 6.59, *p-value* ≤0.001]. In contrast, the average ratings for the 3q29Del group did not significantly differ from 22q11.2Del for positive [*t*(328) = -2.84, *p- value* = 0.01], disorganization [*t*(329) = -1.81, *p-value* = 0.16], or general [*t*(328) = -2.24, *p-value* = 0.06] symptom domains. 22q11.2Del participants exhibited greater negative symptoms than 3q29Del participants [*t*(329) = - 3.75, *p-value* ≤0.001].

To compare group profiles of specific items (Fig. 1), a second set of ANCOVAs were conducted with the same approach. For almost all items, there was a significant effect of diagnostic group on severity (*p-value*s ≤0.003). Comparing 3q29Del participants with HCs revealed more severe ratings on all items (*p*-values ≤0.003), except for sleep disturbance [t(329) = 2.55, *p-value* = 0.01], which was nominally significant. Differences between 3q29Del and 22q11.2Del participants were primarily in negative symptoms. Participants with 22q11.2Del exhibited more severe social anhedonia [*t*(329) = -6.09, *p-value* ≤0.001], avolition [*t*(329) = -4.19, *p-value* ≤0.001], experience of emotions and self [*t*(329) = -4.16, *p-value* ≤0.001], and impaired ideational richness [*t*(329) = -3.10, *p-value* = 0.002]. Lastly, individuals with 22q11.2Del were rated as more impaired in personal hygiene [*t*(329) = -4.27, *p-value* ≤0.001], and as experiencing greater dysphoric mood [*t(*328) = -4.43, *p-value* ≤0.001]. The only group difference in positive symptoms was more severe suspiciousness in 22q11.2Del [*t*(329) = -4.18, *p-value* ≤0.001].

**Figure 1.**
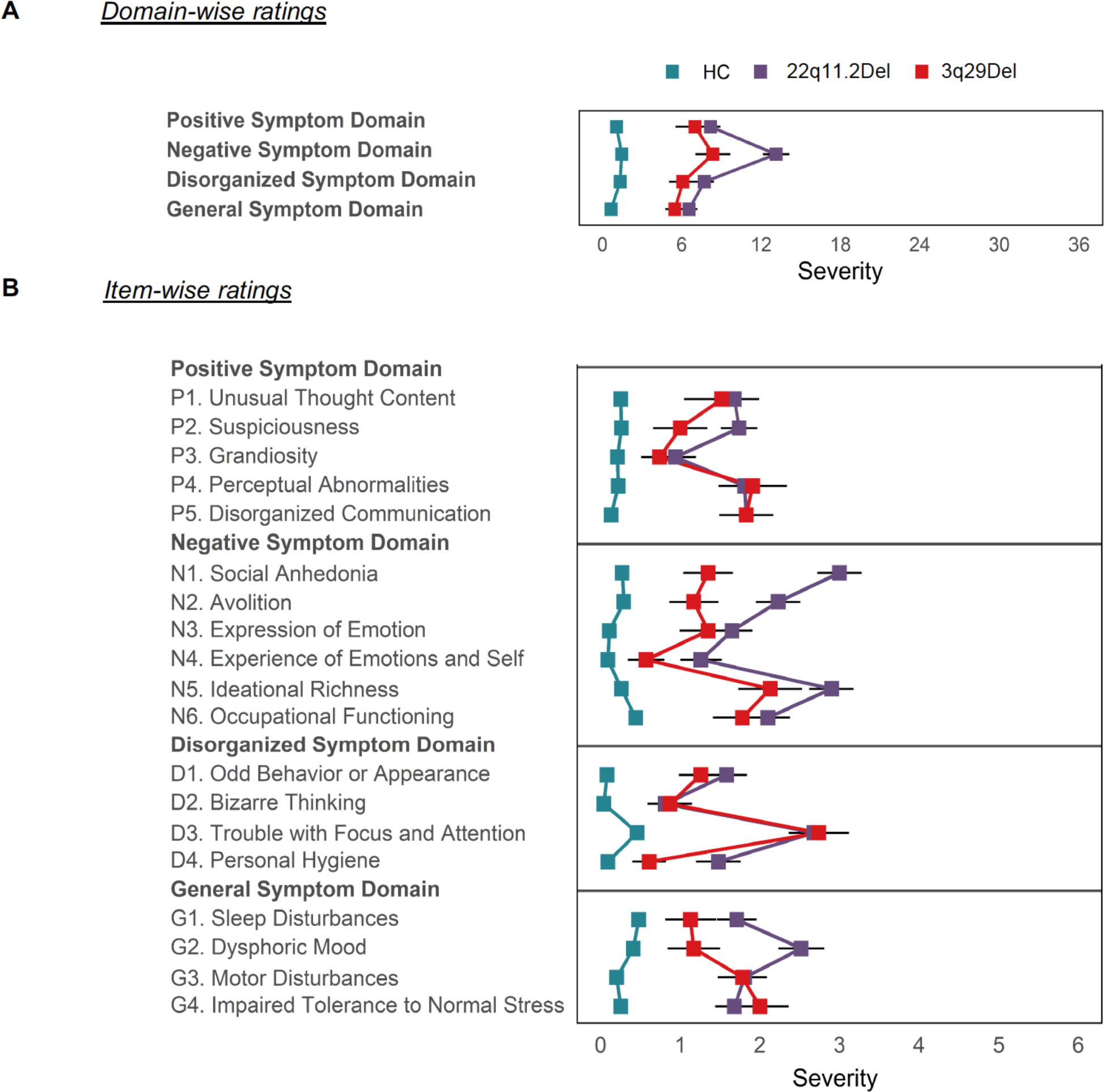
Unadjusted means of individual SIPS ratings for each diagnostic group. Standard error bars are shown. **A)** SIPS domain totals. **B)** Item-specific ratings. For P5 and the positive symptom domain total, 22q11.2Del *N*=30. For G2 and the general symptom domain total, HC *N*=278. *Abbreviations:* Structured Interview for Psychosis-Risk Syndromes, SIPS; healthy controls, HC.

Pairwise comparisons in the complete sample are presented in Table 3, along with untransformed means and standard errors of each group for interpretability on the original scale. See Table S2 for sex-adjusted means and standard errors after log-transformation. The same pattern of results as reported above were found in the smaller sub-sample matched for age and sex. (Tables S3, S4).

### 3.3. Sex differences within groups

We also examined sex differences for the three diagnostic groups (Fig. S3), using ANCOVAs adjusted for age, given no violations in homogeneity of slopes within group. As these analyses were conducted to explore whether there were sex differences in symptoms similar to those observed in research on CHR and psychotic patients, multiple comparisons correction was not made. The differences in domain scores between males and females with 3q29Del were significant only for negative symptoms [*F*(1, 20) = 4.85, *p-value* = 0.04, η^2^p = 0.20], with males exhibiting a more severe profile. Similarly, negative symptoms differed by sex among 22q11.2Del [*F*(1, 28) = 5.10, *p-value* = 0.03, η^2^p = 0.15], and HCs [*F*(1, 276) = 6.94, *p-value* = 0.01, η^2^p = 0.02], with males exhibiting a more severe profile. Lastly, only among HCs, positive symptoms varied between sexes [*F*(1, 276) = 3.93, *p-value* = 0.05, η^2^p = 0.01], with males demonstrating greater severity. Notably, relatively small effect sizes were found in the HC group for both symptom domains.

### 3.4. Structural cerebellar correlates of psychosis-risk in 3q29Del

Prior work by our group using structural MRI acquired from a larger 3q29Del sample (including the scans analyzed herein) has shown that cerebellar hypoplasia and posterior fossa arachnoid cysts are frequently observed among individuals with 3q29Del (Sanchez Russo et al., 2021). We have also shown that 3q29Del participants exhibit smaller cerebellar cortex and larger cerebellar white matter volumes than HCs, and the prevalence of cystic/cyst-like malformations of the posterior fossa is significantly elevated (Sefik et al., 2022).

To determine whether these neuroanatomical abnormalities (see Fig. 2A for a representative image) may be associated with psychotic symptoms among 3q29Del participants, we modeled the dimensional relationship between SIPS ratings for three major symptom domains relevant to psychotic disorders (positive, negative, disorganization) and global and tissue-type specific cerebellar volumes in participants with complete neuroimaging and SIPS data (Table S5). There were no significant differences between left and right hemispheric volumes (Fig. S4), hence bilateral volumes were summed.

**Figure 2.**
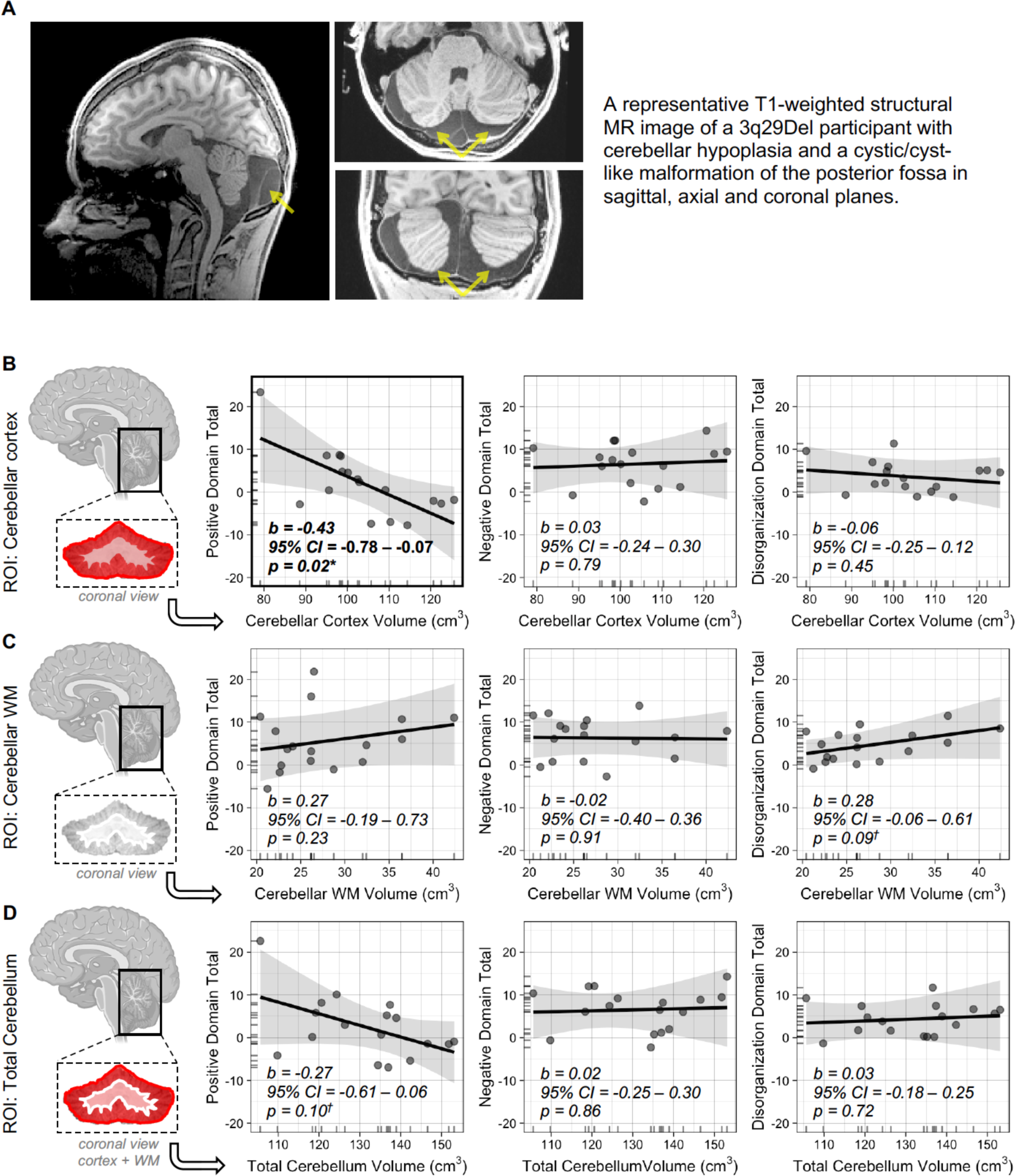
Relationship between cerebellar structure and psychosis-risk symptoms in 3q29Del. A) A representative T1-weighted scan from a 3q29Del participant with cerebellar hypoplasia and a cystic/cyst-like malformation of the posterior fossa (yellow arrow) is shown. **B-D)** Predictor effect plots of the relationship between symptom severity in major SIPS domains relevant to psychotic disorders and global and tissue-type specific cerebellar volumes in 3q29Del (*N*=17). Shaded area is a pointwise confidence band for the fitted values; data points represent partial residuals. *Abbreviations:* region of interest, ROI; white matter, WM; Structured Interview for Psychosis-Risk Syndromes, SIPS.

Results (Fig. 2B-D, Tables S6A-J, Fig. S5) indicate a significant inverse relationship between positive symptom severity and cerebellar cortex volume in 3q29Del participants, correcting for age and sex. Smaller cerebellar cortical volumes were associated with greater positive symptoms (*b* = -0.43, *p-value* = 0.02), and remained significant after eICV correction (*b* = -0.29, *p-value* = 0.03) (Fig. 2B, Tables S6B-C). We also found a trend-level inverse association between positive symptom severity and total cerebellum volume (*b* = -0.27*, p-value* = 0.10) (Fig. 2D, Table S6A), and a trend-level positive association between disorganization symptom severity and cerebellar white matter volume (*b* = -0.28, *p-value* = 0.09) (Fig. 2C, Table S6J).

Given ongoing discussions regarding the merits and limitations of dimensional versus categorical assessment of psychosis (Allardyce et al., 2007; Potuzak et al., 2012), we also took a categorical approach and asked whether the probability of meeting diagnostic criteria for either a psychotic disorder or APSS would recapitulate dimensional results. Findings indicate no significant relationship between interrogated volumes and the likelihood of these outcomes in 3q29Del participants, correcting for age and sex (Tables S7A-C).

Finally, we found no significant association between diagnostic and dimensional indices of psychotic symptoms and the presence of cystic/cyst-like malformations of the posterior fossa in 3q29Del participants (Table S7). There was a trend-level relationship between these radiological findings and the odds of APSS (*p* = 0.08), which may warrant future consideration in a larger sample. See Supplemental Materials for extended results.

## 4. Discussion

Rare pathogenic CNVs that arise from recurrent chromosomal rearrangements are now robustly implicated in psychosis-risk (Malhotra and Sebat, 2012; Rees et al., 2014; Stefansson et al., 2008; Sullivan, 2017), with eight CNV loci surpassing genome-wide significance (Marshall et al., 2017). Among these, 3q29Del has the largest estimated effect size (Marshall et al., 2017; Mulle, 2015), offering a promising opportunity to link a specific genetic mechanism to brain and behavioral phenotypes underlying at least one form of psychosis. Recently, we documented the broader phenotypic spectrum of 3q29Del by direct evaluation of the largest 3q29Del sample reported to date (Murphy et al., 2018; Sanchez Russo et al., 2021). In the present study, we substantially extended these findings by presenting the first in-depth characterization of the profile of psychotic symptoms in 3q29Del and sex- and age-effects on severity. Furthermore, we compared the profiles of 3q29Del to those of 22q11.2Del and one of the largest control samples systematically evaluated with the SIPS. This is particularly important since cross-CNV similarities or differences in symptomology may suggest overlapping or divergent pathogenic mechanisms. Finally, we investigated the neuroanatomical correlates of psychosis-risk in 3q29Del, focusing on macrostructural properties of the cerebellum: a major hindbrain region, whose involvement in higher-order brain functions and psychosis are just beginning to be understood (Morimoto et al., 2021).

First, the present study confirmed that a significant proportion of 3q29Del participants meet criteria for psychosis (17%) or manifest one or more clinically significant attenuated positive symptoms (30%), with 43% meeting the frequency and timing criteria for APSS. Cumulatively, 48% of the 3q29Del sample showed either florid or attenuated psychotic symptoms, as compared to 67% of the 22q11.2Del sample. The rates of psychosis-related phenotypes observed are especially striking considering the young ages of 3q29Del participants. Most of the 3q29Del sample is below the typical age of onset for psychosis prodrome, 15 years-old vs. 17-18 years-old (Addington et al., 2012; Bourgin et al., 2020; Cui et al., 2020). Notably, the prevalence of psychosis or clinically significant attenuated positive symptoms is 67% among 3q29Del individuals aged 17 and older. Given this finding and published rates of transition to psychosis in CHR groups (Fusar-Poli et al., 2012), more transitions to psychosis are to be expected in the present 3q29Del sample.

We note that the 22q11.2Del sample was ascertained through cardiology or immunology clinics, whereas 3q29Del participants were ascertained through a registry. Contrary to the expectation that a clinically ascertained sample may be more severely affected, there were no significant differences in the rates of florid or attenuated psychotic symptoms between 3q29Del and 22q11.2Del samples. 3q29Del participants with a psychotic disorder presented with prodromal symptoms around late-childhood to early-adolescence, consistent with several reports in 22q11.2Del (Chawner et al., 2019; Debbane et al., 2006; Usiskin et al., 1999; Vorstman et al., 2006), although findings vary (Bassett et al., 2003; Ivanov et al., 2003; Murphy et al., 1999). The profile documented in the current sample indicates that early monitoring of psychotic symptoms among individuals with 3q29Del is warranted. Interventions among CHR can delay or prevent the onset of florid psychosis (Preti and Cella, 2010). Thus, early intervention among 3q29Del may also attenuate psychosis severity and improve outcomes.

Further, the relation of sex and age with symptoms in the 3q29Del group was similar to that observed in CHR and psychotic samples (Hafner, 2019; Sullivan et al., 2020; Waford et al., 2015). Males tend to score higher in negative symptoms than females, and both positive and negative symptoms tend to increase with age, although due to the small sample of 3q29Del this correlation was not statistically significant. Studies of both community and clinical samples reveal increases in psychotic-like experiences during adolescence that extends to age ∼19, then begins to decline for those who do not transition to psychosis. The mean age and age-range of the HC group in the present study is in the period when experiences are normatively in decline, hence the inverse relation between age and severity in that group. Given this finding, we performed both age-adjusted and unadjusted analyses to assess the variation in psychotic symptom profiles between groups; both approaches produced the same pattern of results.

Our findings indicate that individuals with 3q29Del are rated significantly higher than HCs on all SIPS domains, while they show remarkable similarity to 22q11.2Del in average ratings for the positive, disorganization, and general symptom domains. However, the 22q11.2Del group exhibited greater negative symptoms than 3q29Del. This parallels an earlier study that compared individuals at CHR with 22q11.2Del to individuals at CHR without 22q11.2Del; the 22q11.2Del group had greater negative symptoms, although the two groups showed comparable positive symptoms (Armando et al., 2012). Individuals with 22q11.2Del also present with greater negative symptoms compared to Williams Syndrome or idiopathic developmental disabilities (Mekori-Domachevsky et al., 2017). In contrast, most individuals with 3q29Del had relatively well-preserved hedonic experiences and social motivation. It is conceivable that the pronounced profile of negative symptoms in 22q11.2Del compared to 3q29Del reflects some level of etiopathogenetic divergence. Disturbances of the brain reward system have been linked to negative symptoms (Barch and Dowd, 2010); hence, reward processing may be an important future direction for cross-comparison studies in these CNVs. Less severe negative symptoms in 3q29Del may also suggest a more favorable profile for response to intervention in psychotic symptoms, considering reports of negative symptoms being more resistant to treatment (Fusar-Poli et al., 2015).

Moreover, our results revealed a significant relationship between positive symptom severity and cerebellar cortex volume in 3q29Del, whereas cystic/cyst-like malformations of the posterior fossa yielded no link with psychotic symptoms. Besides its critical role in sensorimotor coordination, the cerebellum has been suggested to perform a parallel role in higher order functions (Ito, 1993, 2008; Moberget et al., 2014; Schmahmann, 1998), through internal models that, when given its current state, can predict the outcome of an action without performing the action. Accordingly, cerebellar dysfunction has been hypothesized to lead to perceptual abnormalities, such as auditory hallucinations (Ford and Mathalon, 2005; Frith, 2005) and a loss of coherence in speech output (Moberget and Ivry, 2019; Picard et al., 2008), which may reflect dysfunctional predictive mechanisms. Notably, perceptual abnormalities and disorganized communication showed the most prevalent and severe positive symptom ratings in our 3q29Del sample.

The involvement of cerebellar dysfunction in schizophrenia has long been speculated (Andreasen et al., 1998). Emerging findings indicate an increased prevalence of psychosis among patients with cerebellar pathology (Leroi et al., 2002; Liszewski et al., 2004). There are numerous reports of cerebellar abnormalities in idiopathic psychosis (Chen et al., 2013; Gupta et al., 2015; He et al., 2019; Keller et al., 2003; Moberget et al., 2018) and pathology in Purkinje cells, the output neurons of the cerebellar cortex, has been documented in schizophrenia (Maloku et al., 2010; Mavroudis et al., 2017). Further, altered connectivity and morphology of the cerebellum has been identified in CHR groups (Bernard et al., 2017; Cao et al., 2018; Sefik et al., 2023; Smieskova et al., 2010). Notably, a recent study of the Philadelphia Neurodevelopmental Cohort found cerebellar gray matter volume to be a robust predictor of psychotic-like experiences in a large community sample of youths (Moberget et al., 2019), consistent with our findings.

However, how exactly cerebellar abnormalities relate to psychosis remains unclear. The association that we identified between cerebellar cortex volume and positive symptom severity in 3q29Del suggests that one or more genes affected by the hemizygous deletion of this genomic interval may modulate the link between cerebellar development and psychosis-risk in a subset of individuals. In fact, many genes in this interval, including *DLG1* and *BDH1*, show medium-high protein expression in the cerebellum (https://www.proteinatlas.org/). While MRI data were not available for our 22q11.2Del sample, findings in another 22q11.2Del sample also highlight the cerebellum’s significance in the 22q11.2Del neuroendophenotype, with exploratory analyses similarly revealing an association between regional cerebellar cortex volumes and psychotic symptoms in this syndrome (Schmitt et al., 2023). Together with our results, these data suggest a convergence of the two leading genetic risk factors for schizophrenia towards cerebellar connections with psychotic symptoms, which in turn exhibit substantial phenotypic overlap in our comparisons.

Our results also indicate that diagnostic dichotomization does not recapitulate the dimensional brain-behavior relationship identified in 3q29Del. This may be due to reduced power associated with dichotomizing variables. Another relevant issue is the younger mean age of the 3q29Del participants; most have not yet aged through the highest risk period for prodromal or clinical psychosis and may later transition. Additionally, a binary split presupposes a true cutpoint, which may lead to arbitrary divisions of a complex continuum. Our findings support cerebellar cortex morphology as a likely trait marker of psychosis-risk in 3q29Del, which we operationalize as a continuum with varying degrees of severity, distress, and functional impairment.

Our study has several limitations. First, we note that because of the low prevalence of 22q11.2Del and even lower prevalence of 3q29Del, our sample sizes are limited. Enrollment difficulties are bound to occur in rare genetic syndromes, particularly given the significantly elevated risk of psychiatric morbidity. The psychiatric symptoms/syndromes associated with both CNVs also make neuroimaging more challenging. Demands of study participation (e.g., travel to GA) may have barred individuals with more severe presentations from participation; thus, symptom severity could be underestimated. Cerebellar lobes/lobules, and connections with cerebral association cortices as well as the basal ganglia, warrant future investigation in 3q29Del to address questions related to circuitry. Longitudinal follow-up as well as targeted investigations of the cerebellum in the mouse model of 3q29Del (Rutkowski et al., 2021) will be conducted for causal inference and mechanistic insights.

Altogether, our findings indicate that clinical signs of psychosis risk are elevated in 3q29Del participants. Furthermore, our results establish the unique and shared profiles of psychotic symptoms across 3q29Del and 22q11.2Del and highlight cerebellar involvement in elevated psychosis-risk in 3q29Del. Research on these CNVs hold unique promise for elucidating the complex genetic determinants of psychiatric risk; as genotyping technology is improved and more widely accessible, replication in larger samples will become increasingly possible.

## Acknowledgements

We acknowledge the 3q29Del study population, their families and all project members. We thank the Marcus Autism Center for providing clinical assessment resources. We thank all NAPLS2 PIs for their contributions to the HC dataset. Funding for this work was provided in part by National Institute of Mental Health grants R01MH110701, R01MH118534, U01MH081988 and grants by the Robert W. Woodruff Fund and the Predictive Health Initiative of Emory University. This study was not preregistered. The authors have no competing financial interests.

## Data availability

3q29Del data are deposited in the National Institute of Mental Health (NIMH) Data Archive (collection 2614, 3126). Prior to embargo dates, the 3q29Del data and the code used in all statistical analyses are available from the corresponding author, the NAPLS2 data are available from E.F.W. on behalf of the NAPLS consortium, and the 22q11.2Del data are available from O.O. and J.F.C, upon reasonable request.

