## Supplemental Materials for "Psychosis spectrum symptoms among individuals with schizophrenia-associated copy number variants and evidence of cerebellar correlates of symptom severity"

*Table of contents*

**Supplemental figures and tables**……………………………………………………………..……2-23

Figure S1. Age-stratified prevalence rates of florid or attenuated psychotic symptoms in 3q29Del…….2
Table S1. Correlations between SIPS symptom ratings and age at visit, stratified by symptom domain and diagnostic group……………………………..…………………………………………………3
Figure S2. Scatter plots of the relationship between SIPS symptom ratings and age at visit, stratified by symptom domain and diagnostic group.………………………….……………………….…………3
Table S2. Sex-adjusted means and standard errors of SIPS ratings after log-transformation……..……4
Table S3. Demographic and relevant clinical information for age- and sex-matched controls and 22q11.2Del participants and comparison with 3q29Del. ………………………………………………..5
Table S4. Age- and sex-adjusted means and standard errors of SIPS ratings for age- and sex-matched controls and 22q11.2Del participants after log-transformation...............………………………………6
Figure S3. Sex-specific SIPS ratings among deletion groups and HCs…………………………..……….7
Table S5. Demographic, clinical, and MRI-derived volumetric information for the 3q29Del subsample with available neuroimaging and SIPS data…………………………………………………….……….8
Figure S4. Comparison of hemisphere-specific ROI volumes in 3q29Del ………………………….......9
Table S6. Extended linear regression results: Effect of cerebellar volumetric measures on domain-specific SIPS symptom ratings in 3q29Del and polynomial modeling of age………………………10-18
Figure S5. Sensitivity analysis: The relationship between cerebellar cortex volume and positive symptom severity in 3q29Del after removal of an extreme data point……………………..…………..19
Table S7. Extended logistic regression results: Effect of cerebellar volumetric measures on probability of a psychotic disorder or APSS diagnosis in 3q29Del and polynomial modeling of age…………20-22
Table S8. Nested comparison of diagnostic and dimensional phenotypes in 3q29Del participants with versus without posterior fossa arachnoid cyst and mega cisterna magna findings……………………..23

**Supplemental methods**…………………………………………………………………………..…..24-27

Assessment of attenuated and florid psychotic symptoms……………………………………….......24
Cognitive assessment protocols……..………………………………………………..…………..……24-25
Extended methods for structural magnetic resonance imaging……..…………………...…..…………25
Extended statistical analyses…………………………………………………………………………25-26
List of R packages used for statistical analyses………………………………….……………………..26
References for supplemental methods…………………..……………………………….…………...……..27

**Supplemental results**……………………………………………………………………………..…28

Sex differences in SIPS ratings within groups…………………………………………………….......28

**Supplemental figures and tables**

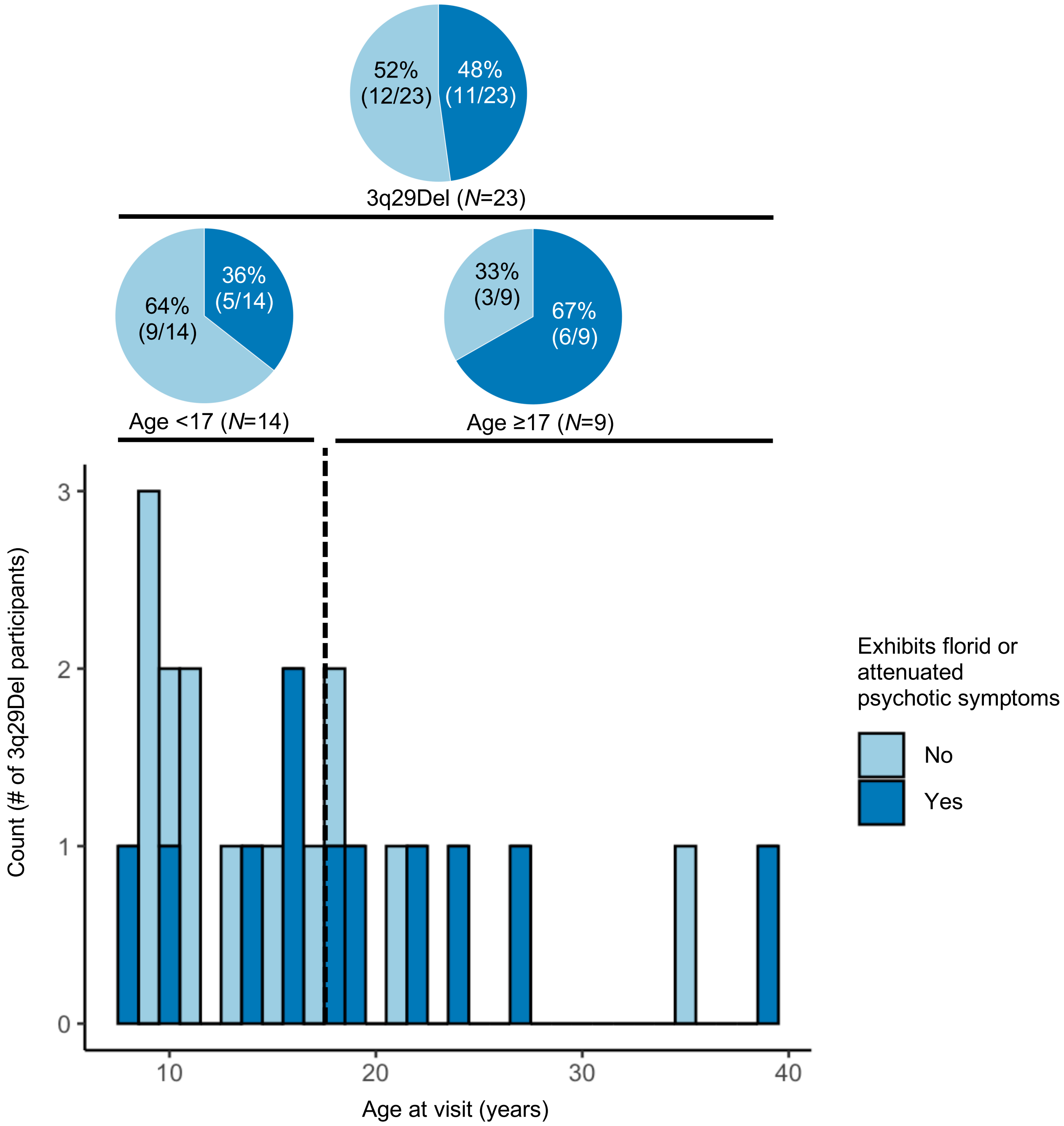

**Figure S1. Age-stratified prevalence rates of florid or attenuated psychotic symptoms in 3q29Del.** 48% of the 3q29Del sample (11/23) meets diagnostic criteria for psychosis or exhibits one or more clinically significant attenuated positive symptoms (i.e., at least one positive symptom rated three or higher on the SIPS). Based on published demographic data on clinical high-risk groups in the general population, the mean age at ascertainment is between 17 and 18 years. Thus, the majority of the 3q29Del participants in the present sample (61%, 14/23) fall below the estimated age for typical onset of both psychosis and the psychosis prodrome (dashed line on histogram marks age 17 years). When stratified by age-group (age <17 years vs age ≥17 years), our results indicate that the prevalence rate of psychosis or clinically significant attenuated positive symptoms among 3q29Del participants younger than age 17 is 36% (5/14), whereas this rate is 67% (6/9) among those aged 17 and older. Too few participants with 3q29Del (*N*=3) were taking antipsychotics to explore treatment effects. *Abbreviations:* Structured Interview for Psychosis-Risk Syndromes, SIPS.

| | Pearson's correlation coefficients between age and symptom ratings | | | Pairwise comparisons of correlation coefficients | |
|---|---|---|---|---|---|
| **SIPS Domains** | **HC** (*N*=279) | **22q11.2Del** (*N*=31) | **3q29Del** (*N*=23) | **3q29Del vs. HC** | **3q29Del vs. 22q11.2Del** |
| Positive Symptom Domain | -0.11[a] | 0.19 | 0.40[a] | z = 2.31<br>*p*-value = 0.02 | z = 0.78<br>*p*-value = 0.43 |
| Negative Symptom Domain | -0.15*[a] | 0.02 | 0.37[a] | z = 2.33<br>*p*-value = 0.02 | z = 1.26<br>*p*-value = 0.21 |
| Disorganization Symptom Domain | -0.08 | 0.15 | 0.15 | z = 1.00<br>*p*-value = 0.32 | z = 0.00<br>*p*-value = 1.00 |
| General Symptom Domain | -0.03 | 0.25 | 0.26 | z = 1.28<br>*p*-value = 0.20 | z = 0.04<br>*p*-value = 0.97 |

**Table S1. Correlations between SIPS symptom ratings and age at visit, stratified by symptom domain and diagnostic group.** To assess the relationship between age and SIPS symptom ratings, Pearson correlations were calculated between age and SIPS domain scores in each study sample. Correlation coefficients were positive and often highest in magnitude for 3q29Del, positive and modest in 22q11.2Del, but negative in HCs. This is consistent with the group differences in the mean and range of age. The 3q29Del group is younger and encompasses an age-range that is typically associated with an increase in symptom severity. The 22q11.2Del and HC groups are older, and the age range in HCs extends into young adulthood when psychotic-like experiences typically decline. Given the group variation in sample size, power for detecting significant correlations between age and symptom severity was lowest in 3q29Del, highest for the HCs, and moderate in 22q11.2Del. Fisher's *r*-to-*Z* transformation was subsequently performed to test the significance of between-group differences in correlation coefficients. The last two columns of the table present results for pairwise comparisons of 3q29Del vs HC, and 3q29Del vs 22q11.2Del. Correlations between age and positive and negative symptom domain ratings showed a significant difference between 3q29Del and HC (*p-values* ≤0.05). *Significant group-specific Pearson correlation (*p-value* ≤0.05). [a]Significant difference between groups in correlation coefficients, based on Fisher's *r*-to-*Z* transformation (*p-value* ≤0.05). For the positive symptom domain total, 22q11.2Del *N*=30. For the general symptom domain total, HC *N*=278. *Abbreviations:* Structured Interview for Psychosis-Risk Syndromes, SIPS; healthy controls, HC.

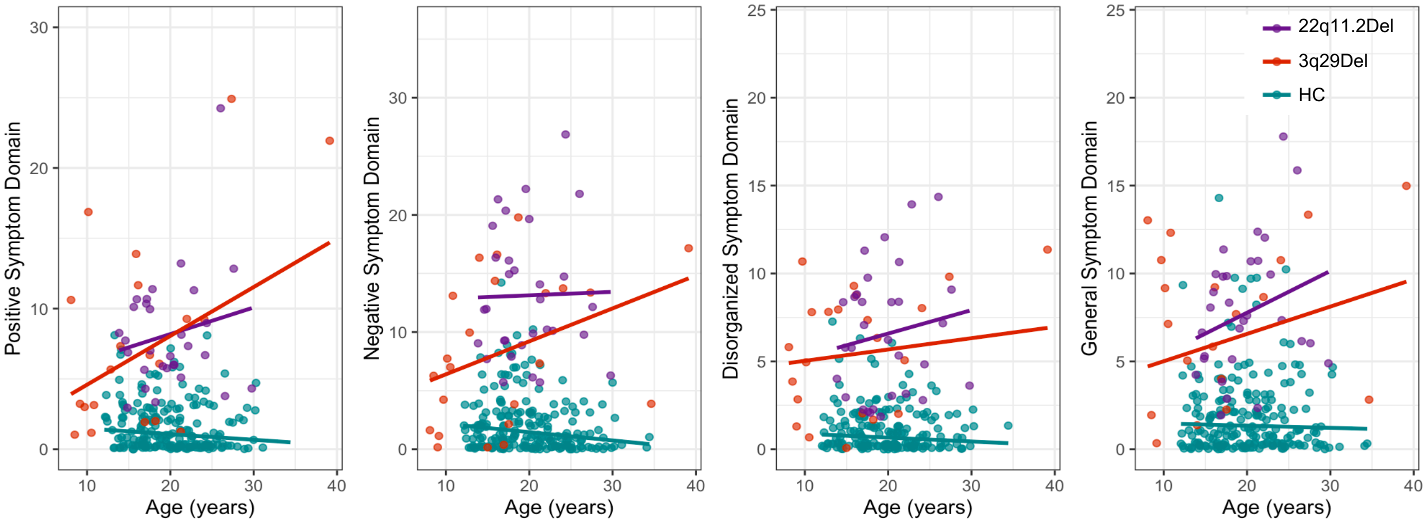

**Figure S2. Scatter plots of the relationship between SIPS symptom ratings and age at visit, stratified by symptom domain and diagnostic group.** The scatterplots above are provided for visual evaluation of the correlation results reported in Table S1. Solid lines represent the line of linear fit for each group. A slight jitter was systematically added to each plot to minimize overplotting. *Abbreviations:* Structured Interview for Psychosis-Risk Syndromes, SIPS; healthy controls, HC.

| SIPS symptom domains and items | HC (*N*=279) | | 22q11.2Del (*N*=31) | | 3q29Del (*N*=23) | |
|---|---|---|---|---|---|---|
| | Adjusted mean | Standard Error | Adjusted mean | Standard Error | Adjusted mean | Standard Error |
| **Positive Symptom Domain** | 0.21 | 0.02 | 0.93 | 0.05 | 0.71 | 0.06 |
| P1. Unusual Thought Content | 0.07 | 0.01 | 0.39 | 0.03 | 0.26 | 0.04 |
| P2. Suspiciousness | 0.07 | 0.01 | 0.39 | 0.03 | 0.20 | 0.03 |
| P3. Grandiosity | 0.05 | 0.01 | 0.21 | 0.03 | 0.16 | 0.03 |
| P4. Perceptual Abnormalities | 0.06 | 0.01 | 0.40 | 0.03 | 0.36 | 0.03 |
| P5. Disorganized Communication | 0.04 | 0.01 | 0.41 | 0.03 | 0.37 | 0.03 |
| **Negative Symptom Domain** | 0.26 | 0.02 | 1.12 | 0.05 | 0.81 | 0.06 |
| N1. Social Anhedonia | 0.07 | 0.01 | 0.56 | 0.03 | 0.29 | 0.03 |
| N2. Avolition | 0.07 | 0.01 | 0.45 | 0.03 | 0.23 | 0.04 |
| N3. Expression of Emotion | 0.03 | 0.01 | 0.35 | 0.03 | 0.27 | 0.03 |
| N4. Experience of Emotion and Self | 0.02 | 0.01 | 0.27 | 0.02 | 0.13 | 0.03 |
| N5. Ideational Richness | 0.07 | 0.01 | 0.54 | 0.03 | 0.40 | 0.04 |
| N6. Occupational Functioning | 0.09 | 0.01 | 0.43 | 0.04 | 0.35 | 0.04 |
| **Disorganization Symptom Domain** | 0.15 | 0.01 | 0.83 | 0.04 | 0.71 | 0.05 |
| D1. Odd Behavior or Appearance | 0.02 | 0.01 | 0.34 | 0.02 | 0.27 | 0.03 |
| D2. Bizarre Thinking | 0.01 | 0.01 | 0.19 | 0.02 | 0.18 | 0.02 |
| D3. Trouble with Focus and Attention | 0.11 | 0.01 | 0.55 | 0.03 | 0.51 | 0.04 |
| D4. Personal Hygiene | 0.02 | 0.01 | 0.30 | 0.02 | 0.14 | 0.03 |
| **General Symptom Domain** | 0.57 | 0.04 | 2.03 | 0.13 | 1.60 | 0.15 |
| G1. Sleep Disturbance | 0.11 | 0.01 | 0.37 | 0.04 | 0.23 | 0.04 |
| G2. Dysphoric Mood | 0.10 | 0.01 | 0.49 | 0.04 | 0.24 | 0.04 |
| G3. Motor Disturbances | 0.05 | 0.01 | 0.38 | 0.03 | 0.37 | 0.03 |
| G4. Impaired Tolerance to Normal Stress | 0.06 | 0.01 | 0.37 | 0.03 | 0.40 | 0.04 |

**Table S2. Sex-adjusted means and standard errors of SIPS ratings after log-transformation.** The log-transformed sex-adjusted means and standard errors for both deletion groups and HCs are reported above. For P5 and the positive symptom domain total, 22q11.2Del *N*=30. For G2 and the general symptom domain total, HC *N*=278. *Abbreviations:* Structured Interview for Psychosis-Risk Syndromes, SIPS; healthy control, HC.

| | 3q29Del (*N*=23) | HC (*N*=69) | 22q11.2Del (*N*=18) | Pairwise comparisons: 3q29Del vs. HC | Pairwise comparisons: 3q29Del vs. 22q11.2Del |
|---|---|---|---|---|---|
| **Age (in years)** | | | | | |
| Mean ± SD | 16.94 ± 8.24 | 17.79 ± 5.84 | 18.70 ± 4.02 | *p-value*[a] = 0.33 | *p-value*[a] = 0.12 |
| Median | 15.89 | 15.89 | 17.57 | | |
| Range | 8.08–39.12 | 12.07–34.41 | 13.85–27.60 | | |
| **Sex, *N (%)*** | | | | | |
| Male | 14 (61%) | 42 (61%) | 14 (78%) | *p-value*[b] = 1.00 | *p-value*[b] = 0.41 |
| Female | 9 (39%) | 27 (39%) | 4 (22%) | | |
| **Race, *N (%)*** | | | | | |
| White | 20 (87%) | 37 (54%) | 7 (78%) | *p-value*[c] = 0.02* | *p-value*[c] = 0.52 |
| Black or African American | 0 (0%) | 11 (16%) | 2 (22%) | | |
| Asian | 0 (0%) | 10 (14%) | 0 (0%) | | |
| More than one race | 3 (13%) | 7 (10%) | 0 (0%) | | |
| Other (i.e., Pacific Islander) | 0 (0%) | 4 (6%) | 0 (0%) | | |
| **Ethnicity, *N (%)*** | | | | | |
| Hispanic/Latino | 1 (4%) | 11 (16%) | 1 (10%) | *p-value*[c] = 0.28 | *p-value*[c] = 0.52 |
| Non-Hispanic/Latino | 22 (96%) | 58 (84%) | 9 (90%) | | |
| **General cognitive abilities** | | | | | |
| Mean ± SD | 73.74 ± 11.98 | 109.90 ± 14.88 | 69.56 ± 14.73 | *p-value*[a] <0.001*** | *p-value*[a] = 0.21 |
| Median | 75 | 111 | 67 | | |
| Range | 46–99 | 72–138 | 46–103 | | |

**Table S3. Demographic and relevant clinical information for age- and sex-matched controls and 22q11.2Del participants and comparison with 3q29Del.** Healthy controls were matched to 3q29Del participants in a 3:1 ratio based on closest available match for sex and age, without replacement. To achieve the closest available match for sex and age between the 22q11.2Del and 3q29Del groups, five females with 22q11.2Del above the age of 19 years were removed from the larger 22q11.2Del sample. For race and ethnicity, 22q11.2Del *N*=9 and *N*=10, respectively. Percentages reflect the fraction of participants with complete data. For general cognitive abilities, HC *N*=67. [a]Wilcoxon rank sum test, [b]Pearson's chi-squared test. [c]Fisher's exact test (due to smaller sub-samples). Two-tailed significance levels: *p-value* ≤0.001'***', *p-value* ≤0.01'**', *p-value* ≤0.05'*', *p-value* ≤0.1'†'. Abbreviations: healthy control, HC; standard deviation, SD.

| Symptom domains and items | HC (*N*=69) | | 22q11.2Del (*N*=18) | | 3q29Del (*N*=23) | | Pairwise comparisons | |
|---|---|---|---|---|---|---|---|---|
| | Adjusted mean | Standard Error | Adjusted mean | Standard Error | Adjusted mean | Standard Error | 3q29Del vs. HC | 3q29Del vs. 22q11.2Del |
| **Positive Symptom Domain** | 0.16 | 0.04 | 0.93 | 0.08 | 0.71 | 0.06 | <0.001 | 0.07 |
| P1. Unusual Thought Content[a] | 0.06 | 0.02 | 0.41 | 0.05 | 0.26 | 0.04 | <0.001 | 0.02 |
| P2. Suspiciousness[a,b] | 0.05 | 0.02 | 0.39 | 0.04 | 0.20 | 0.04 | <0.001 | 0.003 |
| P3. Grandiosity | 0.05 | 0.02 | 0.20 | 0.05 | 0.17 | 0.04 | 0.02 | 0.59 |
| P4. Perceptual Abnormalities[a] | 0.04 | 0.02 | 0.42 | 0.05 | 0.36 | 0.04 | <0.001 | 0.32 |
| P5. Disorganized Communication[a] | 0.02 | 0.02 | 0.36 | 0.04 | 0.37 | 0.04 | <0.001 | 0.77 |
| **Negative Symptom Domain** | 0.23 | 0.04 | 1.14 | 0.07 | 0.81 | 0.06 | <0.001 | 0.003 |
| N1. Social Anhedonia[a,b] | 0.05 | 0.02 | 0.59 | 0.04 | 0.30 | 0.04 | <0.001 | <0.001 |
| N2. Avolition[a,b] | 0.05 | 0.02 | 0.47 | 0.05 | 0.23 | 0.04 | <0.001 | <0.001 |
| N3. Expression of Emotion[a] | 0.02 | 0.02 | 0.42 | 0.04 | 0.27 | 0.04 | <0.001 | 0.01 |
| N4. Experience of Emotion and Self[a,b] | 0.01 | 0.02 | 0.32 | 0.04 | 0.13 | 0.03 | 0.001 | <0.001 |
| N5. Ideational Richness[a] | 0.08 | 0.03 | 0.54 | 0.05 | 0.40 | 0.04 | <0.001 | 0.03 |
| N6. Occupational Functioning[a] | 0.07 | 0.03 | 0.47 | 0.05 | 0.36 | 0.05 | <0.001 | 0.12 |
| **Disorganization Symptom Domain** | 0.13 | 0.03 | 0.85 | 0.06 | 0.71 | 0.05 | <0.001 | 0.17 |
| D1. Odd Behavior or Appearance[a] | 0.00 | 0.02 | 0.35 | 0.04 | 0.28 | 0.03 | <0.001 | 0.17 |
| D2. Bizarre Thinking[a] | -0.01 | 0.02 | 0.24 | 0.04 | 0.19 | 0.03 | <0.001 | 0.23 |
| D3. Trouble with Focus and Attention[a] | 0.10 | 0.02 | 0.53 | 0.05 | 0.50 | 0.03 | <0.001 | 0.68 |
| D4. Personal Hygiene[b] | 0.02 | 0.02 | 0.35 | 0.04 | 0.15 | 0.04 | 0.003 | <0.001 |
| **General Symptom Domain** | 0.22 | 0.04 | 0.89 | 0.07 | 0.69 | 0.07 | <0.001 | 0.10 |
| G1. Sleep Disturbance | 0.10 | 0.03 | 0.38 | 0.05 | 0.23 | 0.05 | 0.02 | 0.03 |
| G2. Dysphoric Mood[a,b] | 0.06 | 0.03 | 0.44 | 0.05 | 0.24 | 0.04 | <0.001 | 0.002 |
| G3. Motor Disturbances[a] | 0.04 | 0.02 | 0.40 | 0.05 | 0.37 | 0.04 | <0.001 | 0.55 |
| G4. Impaired Tolerance to Normal Stress[a] | 0.06 | 0.02 | 0.37 | 0.05 | 0.40 | 0.04 | <0.001 | 0.65 |

**Table S4. Age- and sex-adjusted means and standard errors of SIPS ratings for age- and sex-matched controls and 22q11.2Del participants after log transformation.** SIPS symptom rating comparisons were repeated with age- and sex-matched HCs and 22q11.2Del participants to rule out potential confounding effects of the inter-group age imbalance identified in the larger study sample. '[a]' indicates a significant difference (*p-value* ≤0.01 for domain, *p-value* ≤0.003 for item) between 3q29Del and HC; '[b]' indicates a significant difference (*p-value* ≤0.01 for domain, *p-value* ≤0.003 for item) between 3q29Del and 22q11.2Del. The log-transformed, sex- and age-adjusted means and standard errors for both deletion groups and HCs are reported. *Abbreviations:* Structured Interview for Psychosis-Risk Syndromes, SIPS; healthy control, HC.

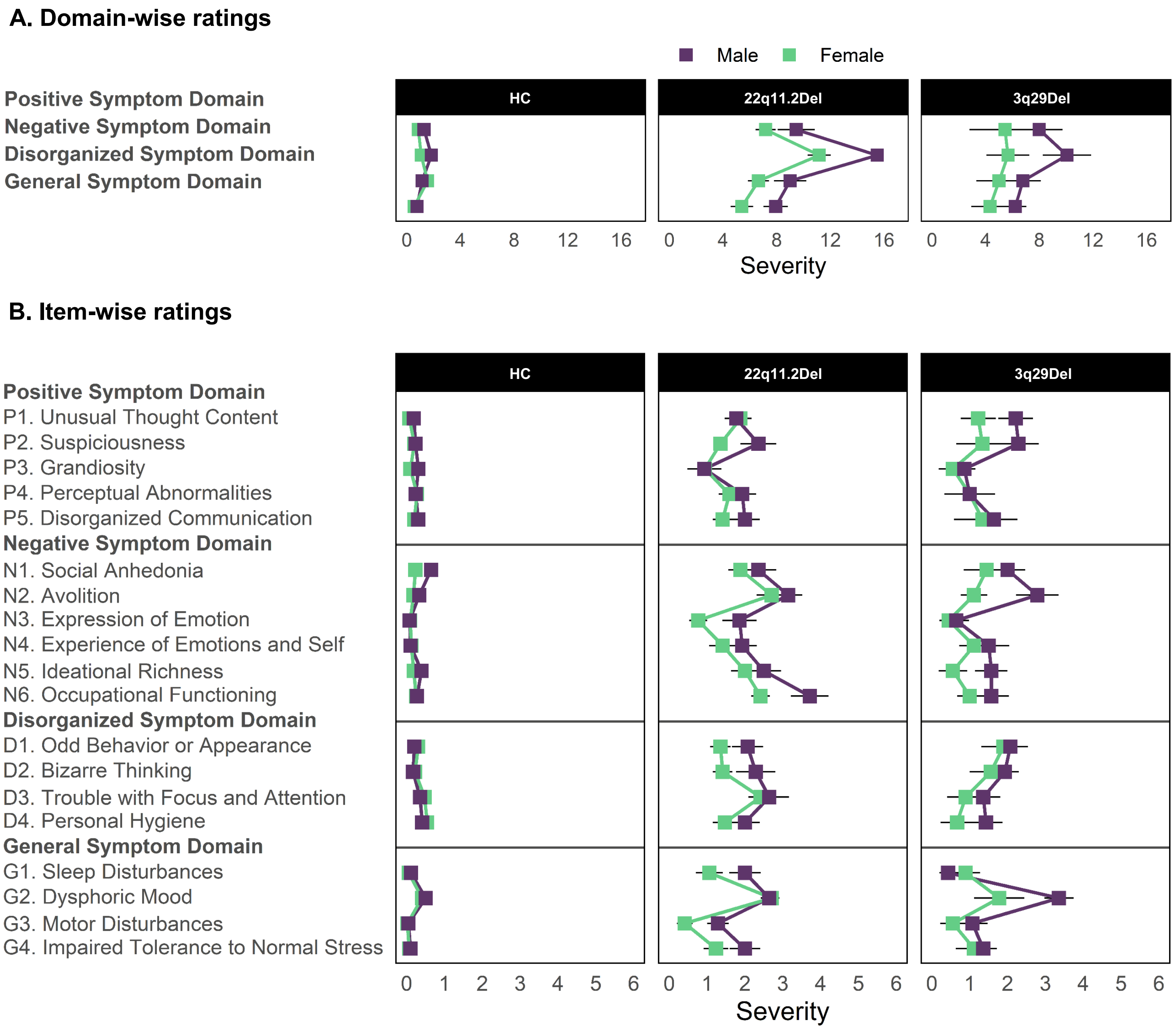

**Figure S3. Sex-specific SIPS ratings among deletion groups and HCs.** Sex differences were investigated in SIPS domain totals (panel **A**) and item-specific SIPS ratings (panel **B**) within each group. Plots reflect the unadjusted means of individual SIPS ratings for each group. Standard error bars are shown. For P5 and the positive symptom domain total, 22q11.2Del *N*=30. For G2 and the general symptom domain total, HC *N*=278. *Abbreviations:* Structured Interview for Psychosis-Risk Syndromes, SIPS; healthy control, HC.

| **3q29Del sub-sample with available neuroimaging and SIPS data** (*N*=17) | |
|---|---|
| *Demographic characteristics* | |
| **Age (in years)** | |
| Mean ± SD | 18.13 ± 9.03 |
| Median | 15.89 |
| Range | 8.08–39.12 |
| **Sex, *N* (%)** | |
| Male | 10 (59%) |
| Female | 7 (41%) |
| **Race, *N (%)*** | |
| White | 15 (88%) |
| More than one race | 2 (12%) |
| **Ethnicity, *N* (%)** | |
| Hispanic/Latino | 1 (6%) |
| Non-Hispanic/Latino | 16 (94%) |
| *Clinical characteristics* | |
| **Psychotic disorder dx., *N (%)*** | 3 (18%) |
| **APSS dx., *N (%)*** | 3 (18%) |
| **Positive symptom domain total, SIPS** | |
| Mean ± SD | 7.65 ± 7.86 |
| Median | 6.00 |
| Range | 0–25 |
| **Negative symptom domain total, SIPS** | |
| Mean ± SD | 8.18 ± 5.53 |
| Median | 7.00 |
| Range | 0–17 |
| **Disorganization symptom domain total, SIPS** | |
| Mean ± SD | 5.41 ± 3.81 |
| Median | 5.00 |
| Range | 0–11 |
| **General cognitive abilities** | |
| Mean ± SD | 71.94 ± 12.02 |
| Median | 73 |
| Range | 46–94 |
| *Neuroimaging measures* | |
| **Total cerebellum volume ($cm^3$)** | |
| Mean ± SD | 131.62 ± 13.82 |
| Median | 135.23 |
| Range | 105.61–153.09 |
| **Cerebellar cortex volume ($cm^3$)** | |
| Mean ± SD | 103.96 ± 12.21 |
| Median | 102.46 |
| Range | 79.11–125.60 |
| **Cerebellar white matter volume ($cm^3$)** | |
| Mean ± SD | 27.66 ± 6.25 |
| Median | 26.18 |
| Range | 20.42–42.37 |
| **Estimated total intracranial volume ($cm^3$)** | |
| Mean ± SD | 1384.30 ± 108.26 |
| Median | 1350.52 |
| Range | 1236.90–1574.65 |

**Table S5. Demographic, clinical, and MRI-derived volumetric information for the 3q29Del subsample with available neuroimaging and SIPS data.** *Abbreviations:* Structured Interview for Psychosis-Risk Syndromes, SIPS; Attenuated psychotic symptom syndrome, APSS; standard deviation, *SD;* diagnosis, dx.

**A.**

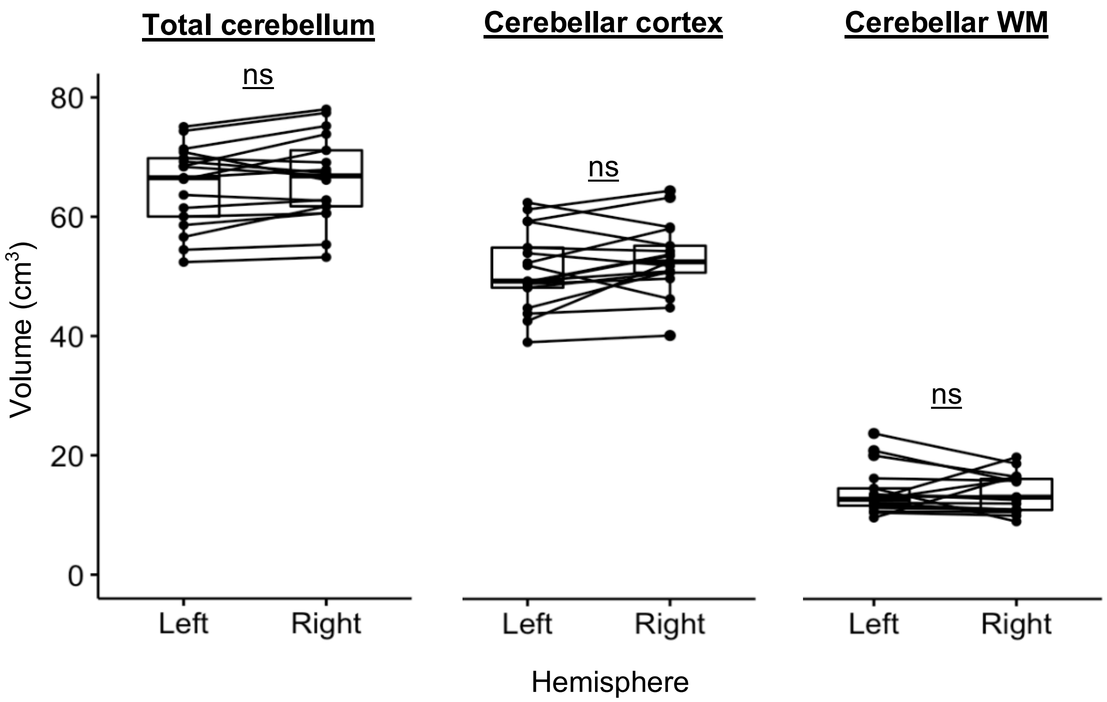

**B.**

| ROI (cm³) | Left Hemisphere | Right Hemisphere | Difference (Left - Right) | *p-value* |
|---|---|---|---|---|
| Total cerebellum volume | | | | |
| Mean ± SD | 65.15 ± 6.86 | 66.46 ± 7.22 | -1.31 ± 2.75 | 0.07 |
| Median | 66.54 | 66.85 | -1.35 | |
| Range | 52.39 – 75.08 | 53.22 – 78.01 | -5.35 – 4.70 | |
| Cerebellar cortex volume | | | | |
| Mean ± SD | 51.14 ± 6.74 | 52.82 ± 6.13 | -1.69 ± 4.11 | 0.11 |
| Median | 49.24 | 52.47 | -1.65 | |
| Range | 38.99 – 62.39 | 40.12 – 64.37 | -9.92 – 5.61 | |
| Cerebellar WM volume | | | | |
| Mean ± SD | 14.02 ± 3.97 | 13.64 ± 3.21 | 0.37 ± 3.60 | 0.13 |
| Median | 12.69 | 13.05 | 0.40 | |
| Range | 9.60 – 23.72 | 8.96 – 19.74 | -7.05 – 5.56 | |

**Figure S4. Comparison of hemisphere-specific ROI volumes in 3q29Del. A)** Box plots showing the paired distribution of left and right hemispheric ROI volumes in 3q29Del (*N*=17). **B)** Descriptive statistics and results of paired-samples tests for inter-hemispheric differences in ROI volumes. In cases where assumptions of paired-samples t-test were violated, the non-parametric Wilcoxon signed-rank test was used. There were no significant volumetric differences between the two hemispheres of any of the ROIs investigated in this study (*p-value* > 0.05). Hence, left and right hemispheric volumes were added to derive a single bilateral volume for each ROI in downstream analyses. *Abbreviations:* Structured Interview for Psychosis-Risk Syndromes, SIPS; magnetic resonance imaging, MRI; region of interest, ROI; white matter, WM; standard deviation, SD; not significant, ns.

**Table S6. Extended linear regression results: Effect of cerebellar volumetric measures on domain-specific SIPS symptom ratings in 3q29Del and polynomial modeling of age.** Multiple linear regression models include sex, age, age$^2$ (when appropriate), and/or eICV (when appropriate) as covariates. Heteroskedasticity-robust Wald tests were performed to sequentially compare simpler models (model 1) to more complex models (model 2) to identify the best fitting polynomial function of age for each ROI. Results indicate that the addition of a quadratic age term in model 2 does not yield a significantly better fit to the data than model 1 (*p-values* > 0.05); hence we favor parsimony and base our statistical inferences on model 1 (highlighted in blue). In **A-J)** heteroskedasticity-robust estimates are reported along with Wald statistics to avoid bias in variance-covariance matrices, given observed violations of statistical assumptions for ordinary least squares regression. In all models, the main effect of cerebellar volume is highlighted in grey for clarity. Schematic illustrations for total cerebellum, cerebellar cortex, and cerebellar white matter are provided below their corresponding regression tables. 3q29Del *N*=17. *Abbreviations:* Structured Interview for Psychosis-Risk Syndromes, SIPS; region of interest, ROI; unstandardized coefficient estimate, *b*; confidence interval, *CI*; degrees of freedom, DF; standard error, *SE;* estimated total intracranial volume, eICV; variance inflation factor, VIF*.* Contrast coding: reference level for the sex variable is female. Two-tailed significance levels: *p-value* ≤0.001‘***’, *p-value* ≤0.01‘**’, *p-value* ≤0.05‘*’, *p-value* ≤0.1‘†’.

**A. Outcome: Positive Symptom Domain Total**

| ***Degree of polynomial*** | ***Explanatory variables*** | ***b*** | ***CI (95%)*** | ***SE b*** | ***p-value*** |
|---|---|---|---|---|---|
| **Linear (Model 1)** **Best-fit** | Intercept | 33.62 | -11.49 – 78.73 | 20.88 | 0.13 |
| | Age (years) | 0.26 | -0.24 – 0.75 | 0.23 | 0.28 |
| | Sex [Male] | 8.88 | 2.48 – 15.27 | 2.96 | 0.01** |
| | **Total Cerebellum Volume ($cm^3$)** | **-0.27** | **-0.61 – 0.06** | **0.16** | **0.10†** |
| | $R^2$ / $R^2$ adjusted | 0.34 / 0.19 | | | |
| | Robust Wald test | F-statistic = 4.02 on 3 and 13 DF, *p-value* = 0.03* | | | |
| **Quadratic (Model 2)** | Intercept | 40.16 | -8.33 – 88.66 | 22.26 | 0.10† |
| | Age | -0.34 | -2.14 – 1.47 | 0.83 | 0.69 |
| | Age$^2$ | 0.01 | -0.03 – 0.06 | 0.02 | 0.51 |
| | Sex [Male] | 8.54 | 2.47 – 14.62 | 2.79 | 0.01** |
| | **Total Cerebellum Volume ($cm^3$)** | **-0.28** | **-0.64 – 0.08** | **0.16** | **0.11** |
| | $R^2$ / $R^2$ adjusted | 0.36 / 0.15 | | | |
| | Robust Wald test | F-statistic = 4.07 on 4 and 12 DF, *p-value* = 0.03* | | | |

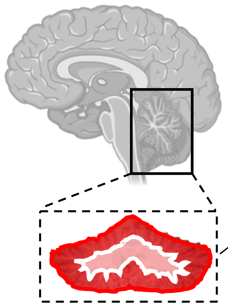

ROI: Total cerebellum volume

Model 1 vs Model 2 – Robust Wald Test:

| | **Residual DF** | **DF** | **F-value** | **Pr(>F)** |
|---|---|---|---|---|
| **1** | 13 | | | |
| **2** | 12 | 1 | 0 | 0.99 |

Final model: Model 1

Table S6 continued.

**B. Outcome: Positive Symptom Domain Total**

| *Degree of polynomial* | *Explanatory variables* | *b* | *CI (95%)* | *SE b* | *p-value* |
|---|---|---|---|---|---|
| **Linear (Model 1) Best-fit** | Intercept | 43.39 | 4.75 – 82.04 | 17.89 | 0.03* |
| | Age (years) | 0.18 | -0.22 – 0.58 | 0.18 | 0.36 |
| | Sex [Male] | 9.70 | 4.21 – 15.19 | 2.54 | 0.002** |
| | **Cerebellar Cortex Volume ($cm^3$)** | **-0.43** | **-0.78 – -0.07** | **0.16** | **0.02*** |
| | $R^2$ / $R^2$ adjusted | 0.49 / 0.37 | | | |
| | Robust Wald test | F-statistic = 11.07 on 3 and 13 DF, *p-value* < 0.001*** | | | |
| **Quadratic (Model 2)** | Intercept | 42.69 | 5.29 – 80.09 | 17.17 | 0.03* |
| | Age | 0.49 | -1.47 – 2.45 | 0.90 | 0.60 |
| | $Age^2$ | -0.01 | -0.05 – 0.04 | 0.02 | 0.74 |
| | Sex [Male] | 10.22 | 3.08 – 17.35 | 3.27 | 0.01** |
| | **Cerebellar Cortex Volume ($cm^3$)** | **-0.45** | **-0.87 – -0.03** | **0.19** | **0.04*** |
| | $R^2$ / $R^2$ adjusted | 0.49 / 0.32 | | | |
| | Robust Wald test | F-statistic = 5.95 on 4 and 12 DF, *p-value* = 0.01** | | | |

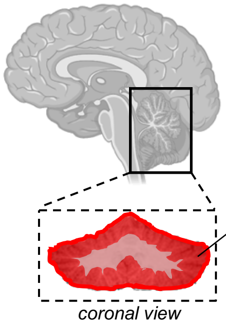

ROI: Cerebellar cortex volume

Model 1 vs Model 2 – Robust Wald Test:

| | Residual DF | DF | F-value | Pr(>F) |
|---|---|---|---|---|
| **1** | 13 | | | |
| **2** | 12 | 1 | 0 | 0.99 |

Final model: Model 1

**C. Outcome: Positive Symptom Domain Total**

| | *Explanatory variables* | *b* | *CI (95%)* | *SE b* | *p-value* | *VIF* |
|---|---|---|---|---|---|---|
| **Secondary model with eICV correction** | Intercept | 70.23 | 31.12 – 109.34 | 17.95 | 0.002** | |
| | Age (years) | 0.21 | -0.23 – 0.64 | 0.20 | 0.32 | 1.20 |
| | Sex [Male] | 10.18 | 5.48 – 14.89 | 2.16 | <0.001*** | 1.43 |
| | eICV ($cm^3$) | -0.03 | -0.05 – -0.01 | 0.01 | 0.01** | 1.37 |
| | **Cerebellar Cortex Volume ($cm^3$)** | **-0.29** | **-0.56 – -0.02** | **0.12** | **0.03*** | **1.99** |
| | $R^2$ / $R^2$ adjusted | 0.61 / 0.48 | | | | |
| | Robust Wald test | F-statistic = 28.78 on 4 and 12 DF, *p-value* <0.001*** | | | | |

*Multicollinearity was assessed in models with eICV by computing variance inflation factor (VIF) scores. VIF > 5 was treated as indicative of a problematic amount of collinearity.*

Table S6 continued.

| D. Outcome: Positive Symptom Domain Total | | | | | |
|---|---|---|---|---|---|
| ***Degree of polynomial*** | ***Explanatory variables*** | ***b*** | ***CI (95%)*** | ***SE b*** | ***p-value*** |
| **Linear (Model 1)** | Intercept | -8.00 | -20.09 – 4.10 | 5.60 | 0.18 |
| | Age (years) | 0.34 | -0.07 – 0.76 | 0.19 | 0.10[†] |
| | Sex [Male] | 3.65 | -4.27 – 11.57 | 3.66 | 0.34 |
| **Best-fit** | **Cerebellar White Matter Volume ($cm^3$)** | **0.27** | **-0.19 – 0.73** | **0.21** | **0.23** |
| | $R^2$ / $R^2$ adjusted | 0.26 / 0.09 | | | |
| | Robust Wald test | F-statistic = 11.58 on 3 and 13 DF, *p-value* <0.001*** | | | |
| **Quadratic (Model 2)** | Intercept | -11.11 | -66.04 – 43.82 | 25.21 | 0.67 |
| | Age | 0.55 | -2.90 – 4.01 | 1.58 | 0.73 |
| | $Age^2$ | -0.00 | -0.08 – 0.07 | 0.04 | 0.89 |
| | Sex [Male] | 3.64 | -4.75 – 12.03 | 3.85 | 0.36 |
| | **Cerebellar White Matter Volume ($cm^3$)** | **0.31** | **-0.70 – 1.33** | **0.47** | **0.51** |
| | $R^2$ / $R^2$ adjusted | 0.26 / 0.01 | | | |
| | Robust Wald test | F-statistic = 7.10 on 4 and 12 DF, *p-value* = 0.004** | | | |

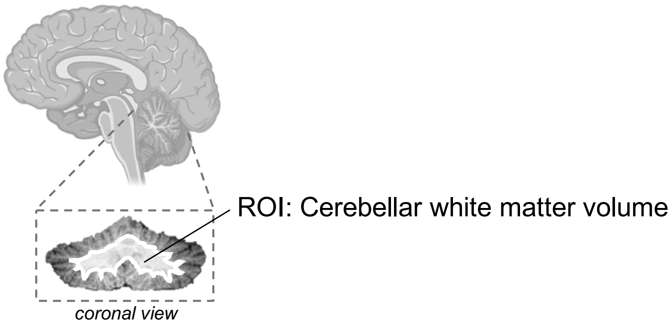

Model 1 vs Model 2 – Robust Wald Test:

| | **Residual DF** | **DF** | **F-value** | **Pr(>F)** |
|---|---|---|---|---|
| **1** | 13 | | | |
| **2** | 12 | 1 | 0 | 0.99 |

Final model: Model 1

Table S6 continued.

| E. Outcome: Negative Symptom Domain Total | | | | | |
|---|---|---|---|---|---|
| ***Degree of polynomial*** | ***Explanatory variables*** | ***b*** | ***CI (95%)*** | ***SE b*** | ***p-value*** |
| **Linear (Model 1)** | Intercept | -1.46 | -36.03 – 33.11 | 16.00 | 0.93 |
| | Age (years) | 0.28 | 0.05 – 0.52 | 0.11 | 0.02* |
| | Sex [Male] | 2.76 | -4.21 – 9.74 | 3.23 | 0.41 |
| **Best-fit** | **Total Cerebellum Volume (cm$^3$)** | **0.02** | **-0.25 – 0.30** | **0.13** | **0.86** |
| | $R^2$ / $R^2$ adjusted | 0.24 / 0.07 | | | |
| | Robust Wald test | F-statistic = 3.90 on 3 and 13 DF, *p-value* = 0.03* | | | |
| **Quadratic (Model 2)** | Intercept | -4.68 | -42.10 – 32.74 | 17.17 | 0.79 |
| | Age | 0.57 | -0.78 – 1.93 | 0.62 | 0.37 |
| | Age$^2$ | -0.01 | -0.04 – 0.02 | 0.01 | 0.66 |
| | Sex [Male] | 2.93 | -4.70 – 10.55 | 3.50 | 0.42 |
| | **Total Cerebellum Volume (cm$^3$)** | **0.03** | **-0.25 – 0.30** | **0.13** | **0.84** |
| | $R^2$ / $R^2$ adjusted | 0.25 / 0.002 | | | |
| | Robust Wald test | F-statistic = 2.39 on 4 and 12 DF, *p-value* = 0.12 | | | |

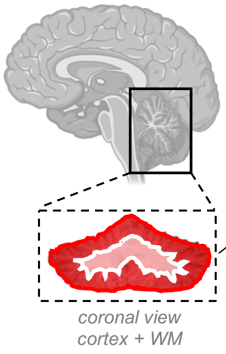

ROI: Total cerebellum volume

Model 1 vs Model 2 – Robust Wald Test:

| | **Residual DF** | **DF** | **F-value** | **Pr(>F)** |
|---|---|---|---|---|
| **1** | 13 | | | |
| **2** | 12 | 1 | 0 | 0.99 |

Final model: Model 1

Table S6 continued.

**F. Outcome: Negative Symptom Domain Total**

| ***Degree of polynomial*** | ***Explanatory variables*** | ***b*** | ***CI (95%)*** | ***SE b*** | ***p-value*** |
|---|---|---|---|---|---|
| **Linear (Model 1)** **Best-fit** | Intercept | -2.15 | -29.68 – 25.37 | 12.74 | 0.87 |
| | Age (years) | 0.29 | 0.04 – 0.54 | 0.12 | 0.03* |
| | Sex [Male] | 2.71 | -3.78 – 9.19 | 3.00 | 0.38 |
| | **Cerebellar Cortex Volume ($cm^3$)** | **0.03** | **-0.24 – 0.30** | **0.13** | **0.79** |
| | $R^2$ / $R^2$ adjusted | 0.24 / 0.07 | | | |
| | Robust Wald test | F-statistic = 3.54 on 3 and 13 DF, *p-value* = 0.05* | | | |
| **Quadratic (Model 2)** | Intercept | -2.69 | -29.62 – 24.24 | 12.36 | 0.83 |
| | Age | 0.53 | -0.97 – 2.03 | 0.69 | 0.46 |
| | $Age^2$ | -0.01 | -0.04 – 0.03 | 0.02 | 0.74 |
| | Sex [Male] | 3.10 | -4.99 – 11.19 | 3.71 | 0.42 |
| | **Cerebellar Cortex Volume ($cm^3$)** | **0.02** | **-0.29 – 0.33** | **0.14** | **0.90** |
| | $R^2$ / $R^2$ adjusted | 0.25 / -0.001 | | | |
| | Robust Wald test | F-statistic = 2.12 on 4 and 12 DF, *p-value* = 0.14 | | | |

Model 1 vs Model 2 – Robust Wald Test:

| | **Residual DF** | **DF** | **F-value** | **Pr(>F)** |
|---|---|---|---|---|
| **1** | 13 | | | |
| **2** | 12 | 1 | 0 | 0.99 |

Final model: Model 1

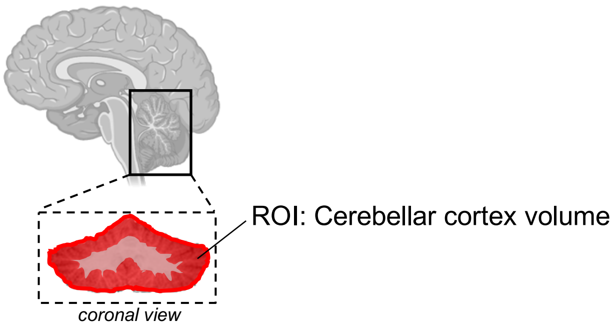

Table S6 continued.

| G. Outcome: Negative Symptom Domain Total | | | | | |
|---|---|---|---|---|---|
| ***Degree of polynomial*** | ***Explanatory variables*** | ***b*** | ***CI (95%)*** | ***SE b*** | ***p-value*** |
| **Linear (Model 1)** **Best-fit** | Intercept | 1.95 | -8.00 – 11.91 | 4.61 | 0.68 |
| | Age (years) | 0.28 | 0.02 – 0.54 | 0.12 | 0.04* |
| | Sex [Male] | 3.19 | -2.15 – 8.53 | 2.47 | 0.22 |
| | **Cerebellar White Matter Volume ($cm^3$)** | **-0.02** | **-0.40 – 0.36** | **0.18** | **0.91** |
| | $R^2$ / $R^2$ adjusted | 0.24 / 0.06 | | | |
| | Robust Wald test | F-statistic = 3.37 on 3 and 13 DF, *p-value* = 0.05* | | | |
| **Quadratic (Model 2)** | Intercept | -4.62 | -42.17 – 32.94 | 17.24 | 0.79 |
| | Age | 0.72 | -1.57 – 3.00 | 1.05 | 0.51 |
| | $Age^2$ | -0.01 | -0.06 – 0.04 | 0.02 | 0.69 |
| | Sex [Male] | 3.17 | -2.52 – 8.87 | 2.62 | 0.25 |
| | **Cerebellar White Matter Volume ($cm^3$)** | **0.08** | **-0.64 – 0.79** | **0.33** | **0.82** |
| | $R^2$ / $R^2$ adjusted | 0.25 / 0.003 | | | |
| | Robust Wald test | F-statistic = 2.14 on 4 and 12 DF, *p-value* = 0.14 | | | |

Model 1 vs Model 2 – Robust Wald Test:

| | **Residual DF** | **DF** | **F-value** | **Pr(>F)** |
|---|---|---|---|---|
| **1** | 13 | | | |
| **2** | 12 | 1 | 0 | 0.99 |

Final model: Model 1

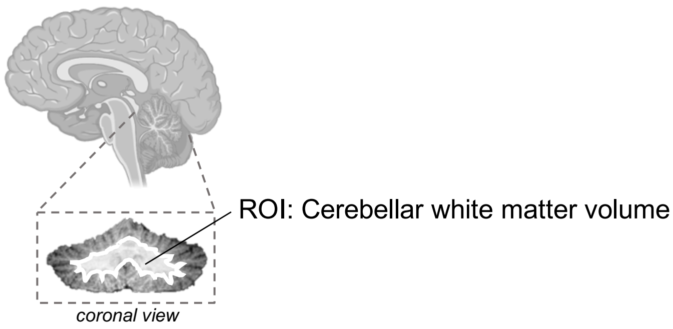

Table S6 continued.

| H. Outcome: Disorganization Symptom Domain Total | | | | | |
|---|---|---|---|---|---|
| ***Degree of polynomial*** | ***Explanatory variables*** | ***b*** | ***CI (95%)*** | ***SE b*** | ***p-value*** |
| **Linear (Model 1)** | Intercept | -1.89 | -26.83 – 23.05 | 11.54 | 0.87 |
| | Age (years) | 0.08 | -0.15 – 0.31 | 0.11 | 0.45 |
| | Sex [Male] | 1.81 | -4.64 – 8.26 | 2.98 | 0.55 |
| **Best-fit** | **Total Cerebellum Volume (cm$^3$)** | **0.04** | **-0.18 – 0.25** | **0.10** | **0.72** |
| | $R^2$ / $R^2$ adjusted | 0.12 / -0.09 | | | |
| | Robust Wald test | F-statistic = 0.70 on 3 and 13 DF, *p-value* = 0.57 | | | |
| **Quadratic (Model 2)** | Intercept | 1.80 | -23.62 – 27.23 | 11.67 | 0.88 |
| | Age | -0.25 | -1.35 – 0.85 | 0.50 | 0.63 |
| | Age$^2$ | 0.01 | -0.02 – 0.03 | 0.01 | 0.50 |
| | Sex [Male] | 1.62 | -5.17 – 8.41 | 3.12 | 0.61 |
| | **Total Cerebellum Volume (cm$^3$)** | **0.03** | **-0.19 – 0.25** | **0.10** | **0.76** |
| | $R^2$ / $R^2$ adjusted | 0.14 / -0.14 | | | |
| | Robust Wald test | F-statistic = 0.91 on 4 and 12 DF, *p-value* = 0.49 | | | |

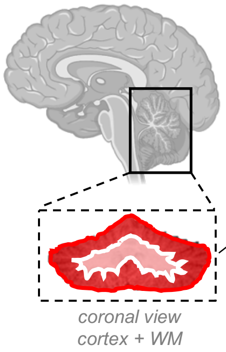

ROI: Total cerebellum volume

Model 1 vs Model 2 – Robust Wald Test:

| | **Residual DF** | **DF** | **F-value** | **Pr(>F)** |
|---|---|---|---|---|
| **1** | 13 | | | |
| **2** | 12 | 1 | 0 | 0.99 |

Final model: Model 1

Table S6 continued.

**I. Outcome: Disorganization Symptom Domain Total**

| *Degree of polynomial* | *Explanatory variables* | *b* | *CI (95%)* | *SE b* | *p-value* |
|---|---|---|---|---|---|
| **Linear (Model 1)** **Best-fit** | Intercept | 9.57 | -10.29 – 29.42 | 9.19 | 0.32 |
| | Age (years) | 0.04 | -0.22 – 0.30 | 0.12 | 0.74 |
| | Sex [Male] | 3.14 | -1.61 – 7.88 | 2.20 | 0.18 |
| | **Cerebellar Cortex Volume ($cm^3$)** | **-0.06** | **-0.25 – 0.12** | **0.08** | **0.45** |
| | $R^2$ / $R^2$ adjusted | 0.13 / -0.06 | | | |
| | Robust Wald test | F-statistic = 0.74 on 3 and 13 DF, *p-value* = 0.55 | | | |
| **Quadratic (Model 2)** | Intercept | 10.14 | -12.14 – 32.41 | 10.22 | 0.34 |
| | Age | -0.21 | -1.58 – 1.16 | 0.63 | 0.74 |
| | $Age^2$ | 0.01 | -0.02 – 0.04 | 0.01 | 0.67 |
| | Sex [Male] | 2.72 | -3.54 – 8.97 | 2.87 | 0.36 |
| | **Cerebellar Cortex Volume ($cm^3$)** | **-0.05** | **-0.29 – 0.19** | **0.11** | **0.68** |
| | $R^2$ / $R^2$ adjusted | 0.15 / -0.13 | | | |
| | Robust Wald test | F-statistic = 1.14 on 4 and 12 DF, *p-value* = 0.39 | | | |

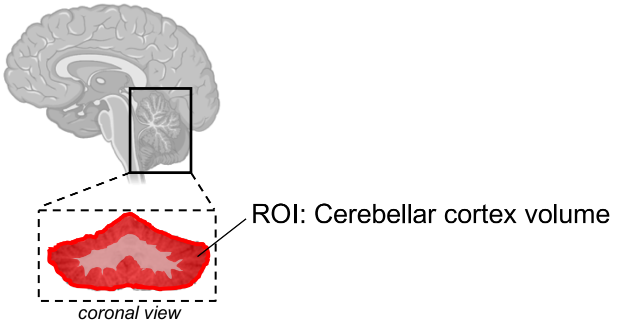

Model 1 vs Model 2 – Robust Wald Test:

| | **Residual DF** | **DF** | **F-value** | **Pr(>F)** |
|---|---|---|---|---|
| **1** | 13 | | | |
| **2** | 12 | 1 | 0 | 0.99 |

Final model: Model 1

Table S6 continued.

| J. Outcome: Disorganization Symptom Domain Total | | | | | |
|---|---|---|---|---|---|
| ***Degree of polynomial*** | ***Explanatory variables*** | ***b*** | ***CI (95%)*** | ***SE b*** | ***p-value*** |
| **Linear (Model 1)** **Best-fit** | Intercept | -3.96 | -10.97 – 3.05 | 3.24 | 0.24 |
| | Age (years) | 0.05 | -0.13 – 0.24 | 0.09 | 0.54 |
| | Sex [Male] | 1.34 | -3.31 – 5.98 | 2.15 | 0.54 |
| | **Cerebellar White Matter Volume (cm$^3$)** | **0.28** | **-0.06 – 0.61** | **0.15** | **0.09†** |
| | $R^2$ / $R^2$ adjusted | 0.29 / 0.13 | | | |
| | Robust Wald test | F-statistic = 6.19 on 3 and 13 DF, *p-value* = 0.01** | | | |
| **Quadratic (Model 2)** | Intercept | -11.07 | -31.63 – 9.49 | 9.44 | 0.26 |
| | Age | 0.53 | -0.68 – 1.75 | 0.56 | 0.36 |
| | Age$^2$ | -0.01 | -0.04 – 0.02 | 0.01 | 0.40 |
| | Sex [Male] | 1.32 | -3.53 – 6.17 | 2.22 | 0.56 |
| | **Cerebellar White Matter Volume (cm$^3$)** | **0.38** | **-0.11 – 0.87** | **0.22** | **0.11** |
| | $R^2$ / $R^2$ adjusted | 0.32 / 0.10 | | | |
| | Robust Wald test | F-statistic = 4.51 on 4 and 12 DF, *p-value* = 0.02* | | | |

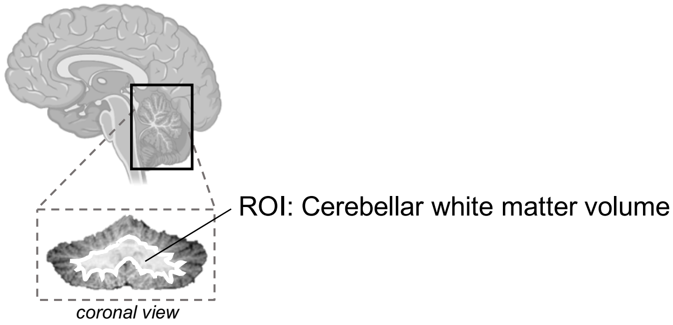

Model 1 vs Model 2 – Robust Wald Test:

| | **Residual DF** | **DF** | **F-value** | **Pr(>F)** |
|---|---|---|---|---|
| **1** | 13 | | | |
| **2** | 12 | 1 | 0 | 0.99 |

Final model: Model 1

**Figure S5. Sensitivity analysis: The relationship between cerebellar cortex volume and positive symptom severity in 3q29Del after removal of an extreme data point. A)** Predictor effect plot showing the relationship between cerebellar cortex volume and SIPS positive symptom ratings (domain total) in *N*=17 3q29Del participants. Predicted values of positive symptom ratings (y-axis) were computed from the multiple linear regression model in Table S5B, while age and sex were held fixed. Visual inspection of this figure suggests that one datapoint indicated by the black arrow (27-year-old female 3q29Del participant) may have high leverage, although it follows the general trend observed in the remaining data. To compute the influence exerted by this single datapoint on the predicted outcome, we calculated its Cook's distance. The mean Cook's distance in the present sample was 0.10, while the Cook's distance for the interrogated participant was 0.52. As values greater than 4 times the mean Cook's distance of the sample may be classified as influential, we performed a sensitivity analysis by removing this datapoint from the sample. Note that this participant is the only female participant who meets diagnostic criteria for psychosis in the 3q29Del neuroimaging dataset**. B)** Multiple linear regression results in *N*=16 3q29Del participants. Heteroskedasticity-robust estimates are reported along with Wald statistics to avoid bias in the variance-covariance matrix, given observed violations of statistical assumptions for ordinary least squares regression. The main effect of cerebellar cortex volume on positive symptom severity is highlighted in grey for clarity. Results of this sensitivity analysis indicate that the coefficient of determination ($R^2$) remains relatively stable after removal of this extreme datapoint. Although statistical power is reduced due to slightly smaller sample size, there was still a significant inverse relationship between cerebellar cortex volume and positive symptom severity, while correcting for age and sex (*p-value* ≤0.05). *Abbreviations:* Structured Interview for Psychosis-Risk Syndromes, SIPS; unstandardized coefficient estimate, *b*; confidence interval, *CI*; degrees of freedom, DF; standard error, *SE.* Contrast coding: reference level for the categorical sex variable is female. Two-tailed significance levels: *p-value* ≤0.001‘***’, *p-value* ≤0.01‘**’, *p-value* ≤0.05‘*’, *p-value* ≤0.1‘†’.

**A**

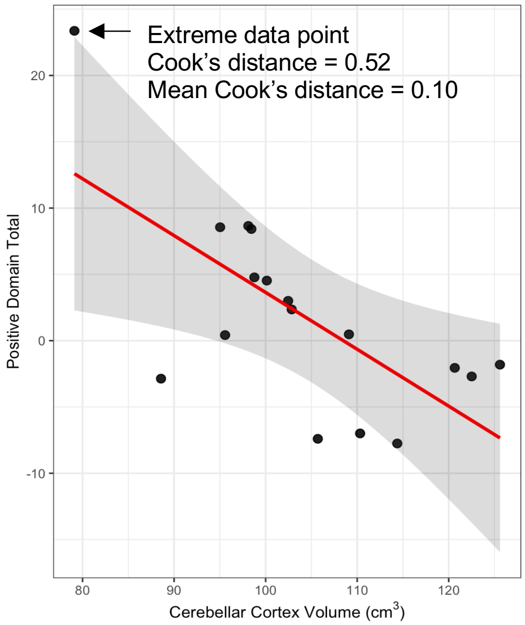

**B**

Regression analysis after removal of one extreme data point:

| **Outcome: Positive Symptom Domain Total** (*N*=16) | | | | |
|---|---|---|---|---|
| ***Explanatory variables*** | ***b*** | ***CI (95%)*** | ***SE b*** | ***p-value*** |
| Intercept | 26.91 | -1.44 – 55.26 | 13.01 | 0.06† |
| Age (years) | 0.15 | -0.24 – 0.54 | 0.18 | 0.42 |
| Sex [Male] | 9.87 | 3.60 – 16.14 | 2.88 | 0.005** |
| **Cerebellar Cortex Volume ($cm^3$)** | **-0.28** | **-0.54 – -0.01** | **0.12** | **0.04*** |
| $R^2$ / $R^2$ adjusted | 0.50 / 0.37 | | | |
| Robust Wald test | F-statistic = 4.86 on 3 and 12 DF, *p-value* = 0.02* | | | |

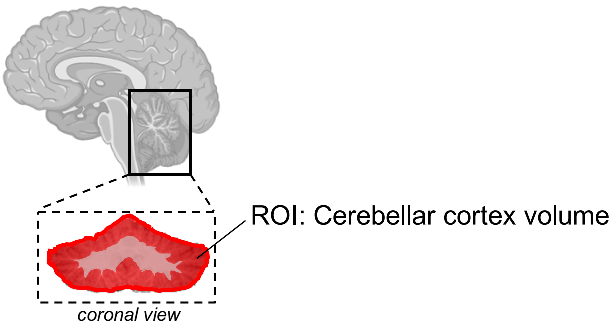

**Table S7. Extended logistic regression results: Effect of cerebellar volumetric measures on probability of a psychotic disorder or APSS diagnosis in 3q29Del and polynomial modeling of age. A-C)** Logistic regression models include sex, age, and/or age$^2$ (when appropriate) as covariates. Analysis of variance (ANOVA) with likelihood ratio test was performed to sequentially compare simpler models (model 1) to more complex models (model 2) to identify the best fitting polynomial function of age for each ROI. Results indicate that the addition of a quadratic age term in model 2 does not yield a significantly better fit to the data than model 1 in any of our analyses (*p-values* > 0.05); hence we favor parsimony and base our statistical inferences on model 1 (highlighted in blue). In all models, the main effect of cerebellar volume is highlighted in gray for clarity. 3q29Del *N*=17. *Abbreviations:* attenuated psychotic symptom syndrome, APSS; region of interest, ROI; odds ratio, *OR*; confidence interval, *CI*; degrees of freedom, DF; standard error, *SE;* diagnosis, dx. Contrast coding: reference level for the sex variable is female. Two-tailed significance levels: p-value ≤0.001‘***’, p-value ≤0.01‘**’, p-value ≤0.05‘*’, p-value ≤0.1‘†’.

**A. Binary outcome: Presence / absence of a psychotic disorder or APSS dx.**

| ***Degree of polynomial*** | ***Explanatory variables*** | ***OR*** | ***CI (95%)*** | ***SE*** | ***p-value*** |
|---|---|---|---|---|---|
| **Linear (Model 1) Best-fit** | Intercept | 2.77 | 0.00 – 4028197.83 | 18.89 | 0.88 |
| | Age (years) | 1.09 | 0.96 – 1.29 | 0.08 | 0.25 |
| | Sex [Male] | 4.34 | 0.24 – 173.78 | 6.86 | 0.35 |
| | **Total Cerebellum Volume (cm$^3$)** | **0.97** | **0.87 – 1.08** | **0.05** | **0.56** |
| | Tjur's $R^2$ | 0.14 | | | |
| **Quadratic (Model 2)** | Intercept | 7.64 | 0.00 – 67707088.23 | 57.13 | 0.79 |
| | Age | 0.97 | 0.50 – 1.90 | 0.32 | 0.93 |
| | Age$^2$ | 1.00 | 0.99 – 1.02 | 0.01 | 0.73 |
| | Sex [Male] | 3.98 | 0.22 – 162.76 | 6.29 | 0.38 |
| | **Total Cerebellum Volume (cm$^3$)** | **0.97** | **0.87 – 1.08** | **0.05** | **0.57** |
| | Tjur's $R^2$ | 0.15 | | | |

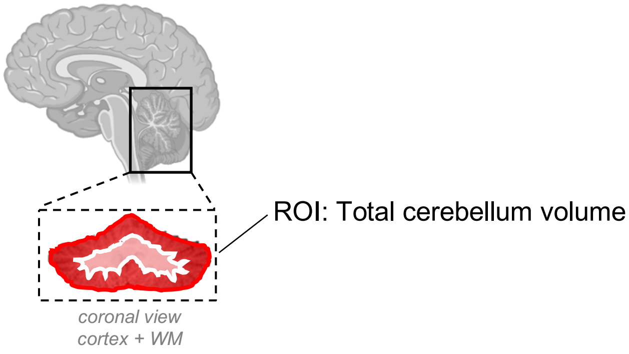

Model 1 vs Model 2 – ANOVA with likelihood ratio test:

| | **Residual DF** | **Residual Deviance** | **DF** | **Deviance** | **Pr(> Chi)** |
|---|---|---|---|---|---|
| **1** | 13 | 19.40 | | | |
| **2** | 12 | 19.28 | 1 | 0.12 | 0.83 |

Final model: Model 1

Table S7 continued.

| B. Binary outcome: Presence / absence of a psychotic disorder or APSS dx. | | | | | |
|---|---|---|---|---|---|
| ***Degree of polynomial*** | ***Explanatory variables*** | ***OR*** | ***CI (95%)*** | ***SE*** | ***p-value*** |
| **Linear (Model 1) Best-fit** | Intercept | 72.82 | 0.00 – 125831051.98 | 458.80 | 0.50 |
| | Age (years) | 1.07 | 0.93 – 1.29 | 0.08 | 0.36 |
| | Sex [Male] | 6.87 | 0.43 – 389.88 | 10.95 | 0.23 |
| | **Cerebellar Cortex Volume ($cm^3$)** | **0.93** | **0.81 – 1.04** | **0.06** | **0.24** |
| | Tjur's $R^2$ | 0.20 | | | |
| **Quadratic (Model 2)** | Intercept | 67.04 | 0.00 – 130020106.90 | 427.82 | 0.51 |
| | Age | 1.10 | 0.54 – 2.35 | 0.39 | 0.78 |
| | $Age^2$ | 1.00 | 0.98 – 1.02 | 0.01 | 0.93 |
| | Sex [Male] | 7.19 | 0.38 – 436.11 | 12.10 | 0.24 |
| | **Cerebellar Cortex Volume ($cm^3$)** | **0.93** | **0.80 – 1.05** | **0.06** | **0.26** |
| | Tjur's $R^2$ | 0.20 | | | |

Model 1 vs Model 2 – ANOVA with likelihood ratio test:

| | **Residual DF** | **Residual Deviance** | **DF** | **Deviance** | **Pr(> Chi)** |
|---|---|---|---|---|---|
| **1** | 13 | 18.24 | | | |
| **2** | 12 | 18.24 | 1 | 0.01 | 0.94 |

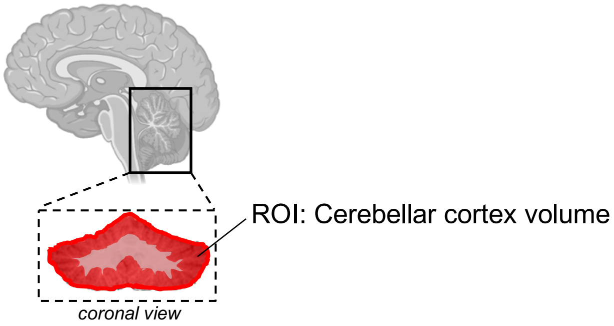

Final model: Model 1

Table S7 continued.

**C. Binary outcome: Presence / absence of a psychotic disorder or APSS dx.**

| ***Degree of polynomial*** | ***Explanatory variables*** | ***OR*** | ***CI (95%)*** | ***SE*** | ***p-value*** |
|---|---|---|---|---|---|
| **Linear (Model 1)** **Best-fit** | Intercept | 0.00 | 0.00 – 1.12 | 0.01 | 0.17 |
| | Age (years) | 1.13 | 0.97 – 1.42 | 0.11 | 0.21 |
| | Sex [Male] | 2.00 | 0.17 – 37.92 | 2.58 | 0.59 |
| | **Cerebellar White Matter Volume ($cm^3$)** | **1.15** | **0.92 – 1.57** | **0.15** | **0.31** |
| | Tjur's $R^2$ | 0.20 | | | |
| **Quadratic (Model 2)** | Intercept | 0.00 | 0.00 – 103.47 | 0.00 | 0.24 |
| | Age | 1.30 | 0.57 – 3.34 | 0.56 | 0.54 |
| | $Age^2$ | 1.00 | 0.98 – 1.02 | 0.01 | 0.73 |
| | Sex [Male] | 1.90 | 0.16 – 35.91 | 2.45 | 0.62 |
| | **Cerebellar White Matter Volume ($cm^3$)** | **1.17** | **0.89 – 1.66** | **0.18** | **0.28** |
| | Tjur's $R^2$ | 0.20 | | | |

Model 1 vs Model 2 – ANOVA with likelihood ratio test:

| | **Residual DF** | **Residual Deviance** | **DF** | **Deviance** | **Pr(> Chi)** |
|---|---|---|---|---|---|
| **1** | 13 | 18.44 | | | |
| **2** | 12 | 18.32 | 1 | 0.12 | 0.73 |

Final model: Model 1

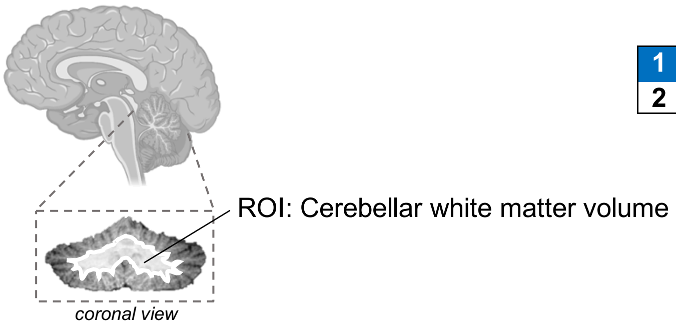

| | 3q29Del sub-sample with available neuroimaging and SIPS data (*N*=17) | | |
|---|---|---|---|
| | **Without PFAC/MCM findings** (*N*=9) | **With PFAC/MCM findings** (*N*=8) | **Test statistics** |
| **Diagnostic evaluation** | | | |
| **Psychosis or APSS dx., N *(%)*** | | | OR = 1.19, |
| Absent | 6 (66.67%) | 5 (62.50%) | 95% CI = 0.11 – 13.47, |
| Present | 3 (33.33%) | 3 (37.50%) | *p-value*[a] = 1 |
| **Psychosis dx., *N (%)*** | | | OR = 0, |
| Absent | 6 (66.67%) | 8 (100%) | 95% CI = 0.00 – 2.54, |
| Present | 3 (33.33%) | 0 (0%) | *p-value*[a] = 0.21 |
| **APSS dx., *N (%)*** | | | OR = INF, |
| Absent | 9 (100%) | 5 (62.50%) | 95% CI = 0.52 – INF, |
| Present | 0 (0%) | 3 (37.50%) | *p-value*[a] = 0.08[†] |
| **Dimensional evaluation** | | | |
| **SIPS Positive Symptom Domain** | | | |
| Mean ± SD | 7.56 ± 9.53 | 7.75 ± 6.11 | *W* = 42.00, |
| Median | 3 | 8 | *p-value*[b] = 0.60 |
| Range | 0 – 25 | 1 – 17 | |
| **SIPS Negative Symptom Domain** | | | |
| Mean ± SD | 7.56 ± 6.21 | 8.88 ± 4.97 | *t* = 0.48, DF = 15, |
| Median | 6 | 7.5 | *p-value*[c] = 0.64 |
| Range | 0 – 17 | 2 – 16 | |
| **SIPS Disorganization Symptom Domain** | | | |
| Mean ± SD | 6.00 ± 4.56 | 4.75 ± 2.92 | *t* = -0.66, DF = 15, |
| Median | 8 | 5 | *p-value*[c] = 0.52 |
| Range | 0 – 11 | 1 – 9 | |

**Table S8. Nested comparison of diagnostic and dimensional phenotypes in 3q29Del participants with versus without posterior fossa arachnoid cyst and mega cisterna magna findings.** [a] Fisher's exact test, [b] Wilcoxon rank sum test, [c] Student's two sample t-test. Results from diagnostic evaluation of psychotic symptoms indicate that there is no statistically significant difference between 3q29Del participants with versus without PFAC/MCM findings in the prevalence of psychotic disorder and/or APSS diagnoses (*p-values* > 0.05). There was a trend-level association (*p-value* ≤0.10) between the odds of meeting diagnostic criteria for APSS and having a PFAC/MCM finding, which may warrant consideration in future studies with larger sample sizes. Results from dimensional evaluation of psychotic symptoms indicate that there was no statistically significant difference between 3q29Del participants with versus without PFAC/MCM findings in SIPS domain totals for positive, negative or disorganization symptoms (*p-values* > 0.05). Non-parametric test statistics are reported in cases where the data do not meet parametric assumptions. Note that given well-established challenges in differentiating PFACs from MCM using conventional MRI (i.e., shared characteristics in appearance), we considered the prevalence rate of these two neuroanatomical findings jointly. *Abbreviations:* diagnosis, dx; posterior fossa arachnoid cyst, PFAC; mega cisterna magna, MCM; attenuated psychotic symptom syndrome, APSS; Structured Interview for Psychosis-Risk Syndromes, SIPS; standard deviation, SD; degrees of freedom, DF; odds ratio, OR; infinity, INF. Two-tailed significance levels: *p-value* ≤0.001‘***’, *p-value* ≤0.01‘**’, *p-value* ≤0.05‘*’, *p-value* ≤0.1‘†’.

### Supplemental methods

#### *Assessment of attenuated and florid psychotic symptoms*

All participants provided written consent to participate in each respective project; in cases when the participant was a minor, a parent or legal guardian provided consent and the minor provided assent. The informed consent process and study procedures were in accordance with Emory University Institutional Review Board for the 3q29Del and 22q11.2Del samples. For NAPLS2, the study procedures were approved by the review boards of each individual site, including Emory University. Participants with 3q29Del were ascertained from the 3q29 registry and between August 2017-February 2020 traveled to Atlanta, GA for deep phenotyping and MRI. The HC sample included 279 individuals from the second cohort of the North American Prodromal Longitudinal Study (NAPLS2), who were evaluated with the SIPS between June 2009-October 2012. Data on 22q11.2Del from the 22q11.2 Clinic in Atlanta, GA were collected between February 2006-June 2007.

In both deletion samples and HCs, the SIPS semi-structured clinical interview was administered by trained personnel, with two main aims: 1) to determine the presence of psychosis, present or past, and 2) to assess the severity of attenuated symptoms (Miller et al., 2003). The 19 items in the SIPS are grouped within four domains: positive, negative, disorganization, and general. Each item is rated on a scale from 0 (*Absent*) to 6 (*Severe and Psychotic* for positive symptoms, *Extreme* for others), with a rating of 3 (*Moderate*) indicating clinical significance. The intensity of experience, any associated distress and impairment, and insight (for positive symptoms) were considered for each rating. Item ratings were summed to produce a total score for each domain. The onset and worsening of symptoms were collected for 3q29Del to determine the presence of a psychosis-risk syndrome. The basis of ratings for 3q29Del and 22q11.2Del also relied on the reports of guardians when present. Overall, the instrument maintains excellent inter-rater reliability (Miller et al., 2003) and is comparable to the performance of another valid instrument in the CHR field (Fusar-Poli et al., 2015). For individuals meeting criteria for psychosis, the Structured Clinical Interview for DSM-5 (SCID-5) (First et al., 2015) or -4 (SCID-4) (First et al., 2004) specified the diagnosis.

#### *Cognitive assessment protocols*

<u>The 3q29Del sample:</u> Participants who were 18-years-old or older completed the Vocabulary, Similarities, Block Design and Matrix Reasoning subtests of the second edition of the Wechsler Abbreviated Scale of Intelligence (WASI-II) (Wechsler, 2011). The scores from all four subsets were then combined to form a Full Score Intelligence Quotient (FSIQ) score to estimate general cognitive ability. Participants who were between seven and 17-years-old completed the Verbal Similarities, Word Definitions, Sequential and Quantitative Reasoning and Matrices subtests of the second edition (school-age form) of the Differential Ability Scales (DAS-II) (Elliot, 2007). Younger participants completed the Verbal Comprehension, Naming Vocabulary, Picture Similarities and Matrices subtests of the second edition (early years form) of the Differential Ability Scales (DAS-II) (Elliot, 2007). The scores from all four subsets were then combined to form a General Conceptual Ability (GCA) composite score to estimate general cognitive ability. See Murphy et al. (2018) for detailed study protocol.

<u>The 22q11.2Del sample</u>**:** Participants who were 17-years-old or older completed the Vocabulary, Arithmetic, Similarities, and Block Design subtests of the third edition of the Wechsler Adult Intelligence Scale (WAIS-III) (Wechsler, 1997). Younger participants completed the equivalent subtests in the third edition of the Wechsler Intelligence Scale for Children (WISC-III) (Wechsler, 1991). As participants completed only a portion of each intelligence scale, an estimation of intellectual functioning hinged upon performance on these subtests. Sattler (2002) identified short-form combinations of individual subtests that could reliably estimate intellectual functioning. The most valid short form for each intelligence scale

was selected for this sample from these combinations; see Appendices A and C in Sattler (2002) for reliability and validity coefficients and conversion tables. For the WISC-III, the best performing short form included the sum of Arithmetic, Vocabulary and Block Design (*r* = 0.88), whereas for the WAIS-III the short form included all four administered subtests (*r* = 0.92). The sum of the scaled scores within each combination were used to derive the estimated full scale deviation quotients (Sattler, 2002).
Healthy controls: All participants were administered two subtests from the first edition of the Wechsler Abbreviated Scale of Intelligence (WASI-I): Vocabulary and Block Design (Wechsler, 1999). The four-subtest WASI was developed from the Wechsler intelligence scales in order to briefly approximate FSIQ, as it relies on its own two- and four-subtest short forms. Two-subtest estimates of FSIQ from the WASI strongly correlate with estimates from the WAIS-III among neurotypical samples (*r* = 0.87) (Weschler, 1999). Performance on these two subtests (Vocabulary and Block Design) were utilized to estimate FSIQ.

#### *Extended methods for structural magnetic resonance imaging (MRI)*

T1- and T2-weighted structural MRI was performed in 17 3q29Del participants with available SIPS data, ages 8.08-39.12 (mean ± SD = 18.13 ± 9.03 years), at the Emory University Center for Systems Imaging Core. Of the six 3q29Del participants who completed SIPS but not MRI, five were medically ineligible and one declined to participate. Images were acquired on a Siemens Magnetom Prisma 3T scanner in the sagittal plane using a 32-channel Prisma head coil with 3D MPRAGE and SPACE sequences, after completion of a training protocol using a mock scanner (i.e., a non-functional replica of the actual MRI machine). In light of previous studies indicating a heightened vulnerability to motion artifacts among children (Brown et al., 2010) and individuals with psychiatric disorders (Pardoe et al., 2016), significant emphasis was placed on completion of the mock scanner session as a prospective motion-reduction technique.

To avoid bias and variability associated with hand-drawn measurements, we performed automatic segmentation of acquired images with the FreeSurfer image analysis pipeline (http://surfer.nmr.mgh.harvard.edu/), using probabilistic information derived from a manually labeled training set (Fischl et al., 2002). For quality control (QC), two trained evaluators (ES, LL) inspected all cerebellar segmentations obtained from 3q29Del participants in axial, sagittal, and coronal reformats to determine whether technical problems (e.g., radiofrequency noise, head coverage, motion artifacts), inconsistencies in tissue boundaries or notable pathology (e.g., arachnoid cysts identified following consultation with a certified neuroradiologist, AEGY) affect segmentation quality. Besides slice-by-slice QC in 2D, the complete cerebellar volume of each participant was additionally visualized using dynamic rotating 3D renderings with the open-source ITK-SNAP software application (http://www.itksnap.org/), to inspect subtle artifacts and variations that might not be immediately apparent in 2D slices. All structural MRI scans included in this study passed 2D and 3D QC.

We highlight that when compared with manual delineation, segmentations obtained by the FreeSurfer software were found by others to yield well-defined boundaries for the cerebellum comparable in accuracy to manual labeling; a previous quantitative evaluation of robustness and accuracy indicated an excellent dice similarity index, as well as high recall and precision values for cerebellar segmentation results acquired by FreeSurfer (Fischl et al., 2002; Lee et al., 2015). Note that, within the present framework, cerebellar cortex refers to the tightly folded outer mantle that is primarily composed of gray matter, while cerebellar white matter refers to the inner core predominantly comprising myelinated nerve fibers.

#### *Extended statistical analyses*

In between-group analyses, since we observed several violations of the statistical assumptions required for ANCOVA (partially due to a likely floor effect in the HC sample), all SIPS scores were log-transformed to improve normality and homogeneity of variances. Age and sex were considered as potential covariates prior to assessing the variation in psychotic symptom profiles between groups. We ran an exploratory Pearson correlation analysis between age and SIPS domain scores separately by diagnostic group and performed Fisher's *r*-to-*Z* transformation to assess the significance of the difference between resulting coefficients. Additionally, we examined sex differences among 3q29Del participants as well as within the comparison groups, using ANCOVAs by symptom domain and then item-wise, while adjusting for age. In sex-specific analyses, assumption violations were observed only in the HC group, hence log-transformation was applied only to HC scores in these analyses. Note that since general symptoms are considered non-specific to psychosis (e.g., impaired tolerance to stress), they were excluded from downstream neuroimaging analyses in the 3q29Del sample.

#### *List of R packages used for statistical analyses*

Pearson's chi-squared tests, Fisher's exact tests, t-tests (paired and unpaired) and Wilcoxon signed-rank tests were performed using the standard R *chisq.test(), fisher.test(), t.test()* and *wilcox.test()* functions. Analyses of variance (ANOVA) and analyses of covariance (ANCOVA) were performed by the standard R *anova()* function. Partial eta-squared was used to assess effect sizes via the *eta_squared()* function from the *effectsize* package (https://CRAN.R-project.org/package=effectsize). Pairwise comparisons for 3q29Del vs. HCs and 3q29Del vs. 22q11.2Del were performed to assess the relative severity of attenuated symptoms using the *glht()* function from the *multcomp* package (https://CRAN.R-project.org/package=multcomp). The standard R *cor.test()* function was used to perform Pearson's correlations. Fisher's *r*-to-*Z* transformation for exploratory correlation analyses between age and symptom domain scores was performed by using the *cocor.indep.groups()* function from the *cocor* package (https://CRAN.R-project.org/package=cocor). The multiple linear regression and binary logistic regression models reported in the neuroimaging arm of the study were performed via the standard R *lm()* and g*lm()* functions (family=binomial, link=logit), respectively. Wald statistics were calculated by the *waldtest()* function from the *lmtest* package (https://CRAN.R-project.org/package=lmtest). Heteroscedasticity-robust estimates were calculated using the *vcovHC()* function from the *sandwich* package (https://CRAN.R-project.org/package=sandwich). Diagnostics plots for linear regression were inspected using the R base function *plot()*. Shapiro-Wilk tests were performed using the standard R *shapiro.test()* function to check for assumptions of normality. Breusch-Pagan and Levene's test were performed using the *bptest()* function from the *lmtest* package (https://CRAN.R-project.org/package=lmtest) and the *levene_test()* function from the *rstatix* package (https://CRAN.R-project.org/package=rstatix) to check for homogeneity of variances, respectively. Graphics were generated by the *ggplot2* (https://CRAN.R-project.org/package=ggplot2)*, ggpubr (https://cran.r-project.org/package=ggpubr)* and *jtools* (https://cran.r-project.org/package=jtools) packages.

***References for supplemental methods***

Brown TT, Kuperman JM, Erhart M, et al. (2010): Prospective motion correction of high-resolution magnetic resonance imaging data in children. *Neuroimage* 53(1):139-145.

Elliott CD (2007): *Manual for the Differential Abilities Scales, 2nd ed*. San Antonio, TX: Harcourt Assessment.

First MB, Williams JB, Karg RS, Spitzer RL (2015): *Structured clinical interview for DSM-5—Research version (SCID-5 for DSM-5, research version; SCID-5-RV).* Arlington, VA: American Psychiatric Association.1-94.

First MB, Gibbon M (2004): *The Structured Clinical Interview for DSM-IV Axis I Disorders (SCID-I) and the Structured Clinical Interview for DSM-IV Axis II Disorders (SCID-II).* in Comprehensive handbook of psychological assessment, Vol 2: Personality assessment. Hoboken, NJ: John Wiley & Sons, Inc. 134-143.

Fischl B, Salat DH, Busa E, Albert M, Dieterich M, Haselgrove C et al. (2002): Whole brain segmentation: automated labeling of neuroanatomical structures in the human brain. *Neuron* 33(3): 341-355.

Fusar-Poli P, Cappucciati M, Rutigliano G, Schultze-Lutter F, Bonoldi I, Borgwardt S, et al. (2015): At risk or not at risk? A meta-analysis of the prognostic accuracy of psychometric interviews for psychosis prediction. *World Psychiatry*. 14:322-332.

Lee DK, Yoon U, Kwak K, Lee JM (2015): Automated Segmentation of Cerebellum Using Brain Mask and Partial Volume Estimation Map. *Comput Math Methods Med* 2015: 167489.

Miller TJ, McGlashan TH, Rosen JL, Cadenhead K, Cannon T, Ventura J, et al. (2003): Prodromal assessment with the structured interview for prodromal syndromes and the scale of prodromal symptoms: predictive validity, interrater reliability, and training to reliability. *Schizophr Bull*. 29:703-715.

Murphy MM, Lindsey Burrell T, Cubells JF, Espana RA, Gambello MJ, Goines KCB, et al. (2018): Study protocol for The Emory 3q29 Project: evaluation of neurodevelopmental, psychiatric, and medical symptoms in 3q29 deletion syndrome. *BMC Psychiatry* 18:183.

Pardoe HR, Kucharsky Hiess R, Kuzniecky R (2016): Motion and morphometry in clinical and nonclinical populations. *NeuroImage 135*: 177–185.

Sattler JM (2002): *Assessment of children: Cognitive applications*. San Diego, CA: Jerome M. Sattler, Publisher, Inc.

Wechsler D (1991): *Manual for the Wechsler Intelligence Scale for Children-III*. San Antonio, TX: The Psychological Corporation.

Wechsler D (1997): *Manual for the Wechsler Adult Intelligence Scale-III*. San Antonio, TX: The Psychological Corporation.

Wechsler D (1999): *Manual for the Wechsler Abbreviated Scale of Intelligence*. San Antonio, TX: The Psychological Corporation.

Wechsler D (2011): *Manual for the Wechsler Abbreviated Scale of Intelligence*, *2nd ed.* Bloomington, MN: Pearson.

### Supplemental results

#### *Sex differences in SIPS ratings within groups*

We examined within-group sex differences for the three diagnostic groups. Statistical analyses included a set of ANCOVAs adjusted for age; values for HCs were log-transformed given assumption violations within this group. As these analyses were conducted to explore whether there were sex differences in symptoms similar to those observed in research on CHR and psychotic patients, correction for multiple comparisons was not made. The unadjusted means and standard errors for each group are presented in Fig. S3.

The differences in domain scores between males and females with 3q29Del were nonsignificant for positive [$F(1,20) = 1.60$, $p = 0.22$], disorganization [$F(1,20) = 1.90$, $p = 0.18$], and general [$F(1,20) = 1.13$, $p = 0.30$] symptoms. Sex did explain variation in negative symptoms [$F(1, 20) = 4.85$, $p = 0.04$, $\eta^2_p = 0.20$], as males exhibited more severe symptoms (age-adjusted mean ± *SD* = 10.42 ± 5.58) than females (age-adjusted mean ± *SD* = 5.12 ± 5.61). Similarly, among the 22q11.2Del group, there was no sex difference in positive [$F(1, 27) = 2.40$, $p = 0.13$], disorganization [$F(1, 28) = 4.19$, $p = 0.05$], or general [$F(1, 28) = 3.07$, $p = 0.09$] symptoms. Only negative symptoms differed by sex [$F(1, 28) = 5.10$, $p = 0.03$, $\eta^2_p = 0.15$], with males with 22q11.2Del exhibiting more severe symptoms (age-adjusted mean ± *SD* = 15.50 ± 5.31) than females (age-adjusted mean ± *SD* = 11.20 ± 5.32). Sex differences among HCs evidenced a similar pattern, with no sex differences for disorganization [$F(1, 276) = 1.21$, $p = 0.27$], or general [$F(1, 276) = 2.96$, $p = 0.09$] symptoms, and negative symptoms were rated higher in males (log-transformed age-adjusted mean ± *SD* = 0.311 ± 3.08) than females (log-transformed age-adjusted mean ± *SD* = 0.21 ± 3.10) [$F(1, 276) = 6.94$, $p = 0.01$, $\eta^2_p = 0.02$]. However, sex differences among HCs were observed in positive symptoms [$F(1, 276) = 3.93$, $p = 0.05$, $\eta^2_p = 0.01$], with greater severity among males (log-transformed age-adjusted mean ± *SD* = 0.25 ± 2.79) versus females (log-transformed age-adjusted mean ± *SD* = 0.18 ± 2.78).

Sex differences were also tested in item-wise ratings within each group, while adjusting for age. For 3q29Del, only one item, impaired ideational richness, marginally varied by sex, with males rated higher [$F(1,20) = 4.71$, $p = 0.04$, $\eta^2_p = 0.19$]. In contrast, males were rated significantly higher than females with 22q11.2Del on ratings for suspiciousness [$F(1,20) = 4.32$, p = 0.05, $\eta^2_p = 0.13$], social anhedonia [$F(1,28) = 6.06$, $p = 0.02$, $\eta^2_p = 0.18$], experience of emotion and self [$F(1,20) = 5.43$, $p = 0.03$, $\eta^2_p = 0.16$], and bizarre thinking [$F(1,20) = 6.22$, $p = 0.02$, $\eta^2_p = 0.18$]. Lastly, for HCs, males were rated higher than females on grandiosity [$F(1,276) = 10.27$, $p = 0.001$, $\eta^2_p = 0.04$], disorganized communication [$F(1,276) = 6.02$, $p = 0.01$, $\eta^2_p = 0.02$], and occupational functioning [$F(1,276) = 7.77$, $p = 0.014$, $\eta^2_p = 0.03$].
